# Modelling the potential spread of Clade Ib MPXV in Asian cities

**DOI:** 10.1101/2024.10.16.24315640

**Authors:** Shihui Jin, Gregory Gan, Akira Endo, Kiesha Prem, Rayner Kay Jin Tan, Jue Tao Lim, Keisuke Ejima, Borame L Dickens

## Abstract

**Background:** The ongoing 2023–2024 mpox outbreak in several African countries, driven by the novel Clade Ib strain, has resulted in imported cases reported in Sweden, Thailand, and India. The potentially high transmissibility of this new strain and shifts in transmission modes may make territories in Asia, which were minimally affected by previous mpox waves, susceptible to community-wide transmission following importation. While this highlights the importance of early preparedness, current knowledge of the virus’s transmission dynamics remains too limited to effectively inform policy-making and resource planning.

**Methods:** A compartmental model was constructed to characterise the potential mpox transmission dynamics. Importation-triggered outbreaks were simulated in 37 Asian cities under scenarios with one, three, and five initial local infections. The impacts of various non-pharmaceutical interventions (NPIs), including isolation and quarantine, were projected and compared.

**Findings:** Our simulations revealed substantial disparities in outbreak sizes among the 37 Asian cities, with large-scale outbreaks expected in territories with a high proportion of sexually active individuals at risk or low immunity from smallpox vaccination. Total case count in one year following initial local infections would increase linearly with initial infection size. In the scenario with three initial local infections, up to 340 cases per million residents were expected without interventions. Isolation for diagnosed cases was projected to lower the outbreak size by 43.8% (interquartile range [IQR]: 42.7%–44.5%), 67.8% (IQR: 66.5%–68.9%), 80.8% (IQR: 79.5%–82.0%), and 88.0% (IQR: 86.8%–89.1%) when it reduced interpersonal contacts by 25%, 50%, 75%, and 100%, respectively. Quarantining close contacts would contribute to a further decrease in cases of up to 22 percentage points over one year.

**Interpretation:** The potential mpox outbreak in an Asian setting could be alleviated through strong surveillance and timely response from stakeholders. NPIs are recommended for outbreak management due to their demonstrated effectiveness and practicability.

**KEY MESSAGES:** *What is already known on this topic:* Clade Ib monkeypox virus (MPXV) is circulating in the Democratic Republic of the Congo (DRC) and its neighbouring countries, with imported cases being identified globally. Meanwhile, evidence indicates that the virus can be transmitted through both sexual and non-sexual routes, raising concerns about its potential spread in the general population. To prevent a global outbreak, the World Health Organization (WHO) suggested countries around the globe to prepare in advance.

*What this study adds:* Our simulations quantified the potential disease burden of an mpox outbreak triggered by a once-time importation event in 37 major Asian cities with varying pre-existing immunity levels and populations at higher risk due to frequent sexual activities. The compartmental modelling framework developed in this study also projected the effectiveness of diverse non-pharmaceutical intervention (NPI) strategies in outbreak control, providing policy-makers with guidance for effective public health crisis management.

*How this study might affect research, practice or policy:* An importation-triggered mpox outbreak can be substantially mitigated with powerful disease surveillance and a prompt response of the stakeholders, but may also lead to severe consequences with high morbidity and mortality if not addressed in time in cities with a large highly sexually active population. Various NPIs, particularly isolating infected cases, are recommended for curbing the disease outbreak due to their feasibility and effectiveness in the Asian setting.

## 1. INTRODUCTION

An outbreak caused by Clade I monkeypox virus (MPXV) was estimated to have emerged in September 2023 in South Kivu province [1], Democratic Republic of the Congo (DRC), with transmission likely driven primarily by sexual contact. The novel strain of the outbreak, later designated as Clade Ib, resulted in surging cases reported in the DRC, alongside the endemic Clade Ia already circulating in other provinces, and quickly spread to neighbouring African countries, including Burundi, Rwanda, Uganda, and Kenya. As of September 2024, importation has also been detected outside the African continent, in Sweden, Thailand, and India [2,3]. In response to the rapid spread of Clade Ib in Africa, the World Health Organisation re-declared mpox as a public health emergency of international concern (PHEIC), aiming to raise the global awareness and prevent a repeat of the 2022 mpox outbreak, whose global circulation could potentially be attributed to initial negligence in surveillance and prevention [4].

During the 2022 wave caused by Clade IIb MPXV, over 90% of the cases reported were from Europe and the Americas. In contrast, the Western Pacific, Southeast Asia, and Eastern Mediterranean regions—encompassing most Asian countries—collectively accounted for fewer than 5% of all Clade IIb cases [5]. Whilst the possibility of high under-ascertainment exists due to the limited surveillance efforts and reluctance in seeking medical help due to stigmas associated with the infection [6], the proportion of the population in Asia with active infection-induced immunity against the virus is still likely to be much lower compared to other regions of the world with higher cumulative prevalence.

While Clade IIb MPXV was predominantly spread through sexual transmission within the gay, bisexual and other men-who-have-sex-with-men (GBMSM) community [7], heterosexual contact appears to play a more critical role in the ongoing 2023–2024 mpox outbreak driven by Clade Ib in the DRC and neighbouring countries, as female cases, especially sexual workers, comprised approximately half of all the confirmed infections [1,8]. Apart from the well-documented sexual mode of transmission, evidence from the ongoing outbreak, including the non-trivial contribution of children to reported case counts, has suggested potential higher prevalence of non-sexual transmission routes compared to the previous 2022 outbreak [8–10]. These two possible changes in transmission modes, together with the high global connectivity and international travel, raise the risk of an importation-triggered mpox outbreak affecting the general population, particularly in an Asia setting with minimal prior exposure to the virus.

Throughout the COVID-19 pandemic, territories in Asia gained valuable experience in implementing non-pharmaceutical interventions (NPIs) [11,12]. Social distancing measures like quarantine and isolation, which reduced interpersonal contact, effectively contained the disease transmission [13]. These successful stories offer hope for the authorities in this area to leverage NPIs to curb a potential mpox outbreak, should it be widespread in the general community. Furthermore, the absence of definitive evidence for respiratory transmission [14] for MPXV implies that effective outbreak management might be achieved through less stringent NPIs, such as isolating infected individuals, compared to those implemented during the COVID-19 pandemic.

Nevertheless, few NPIs were applied to manage the 2022 mpox outbreak, which affected only a limited subpopulation. This creates great uncertainty about their true effectiveness and sufficiency in averting a more widespread mpox outbreak once it establishes itself in the general population. In addition, the distinct transmission patterns of Clade Ib MPXV, especially regarding the prolonged infectious duration and the combination of sustained transmission through sexual and non-sexual routes [10,15], make it challenging to directly extrapolate the impacts of these containment measures from COVID-19 surveillance data.

To address these gaps, we performed this study to model potential mpox outbreaks in 37 Asian cities, covering all key territories in this region. We collected territory-specific data on the immunity levels from smallpox vaccination, age structure, and sexually active populations which were assumed of higher transmission risk. These statistics enabled us to simulate importation-triggered outbreaks utilising a modified deterministic susceptible-exposed-infectious-recovered (SEIR) model and the transmissibility parameters estimated from the surveillance data in the DRC. We further projected the impacts of NPIs on curbing outbreaks and reducing disease burden. These results collectively provide insights into factors shaping the outbreak dynamics, informing policy-makers of strategies to pre-empt or mitigate mpox transmission across the continent.

## 2. METHODS

### 2.1 SEIR-style transmission model

We adapted a deterministic SEIR compartmental model to characterise potential disease transmission patterns, as shown in Figure 1. Additional compartments, including Quarantined (*Q*), Diagnosed (*C*), and Dying (*D*), were incorporated into the model to facilitate a more accurate reflection of a real-world outbreak, with disease surveillance, variations in disease burden among individuals, and the probable presence of intervention measures. Nevertheless, the number of individuals transitioning to Compartment *Q* from the other compartments was calculated separately at each time step, rather than being solved simultaneously with the differential equation system.

**Figure 1.**
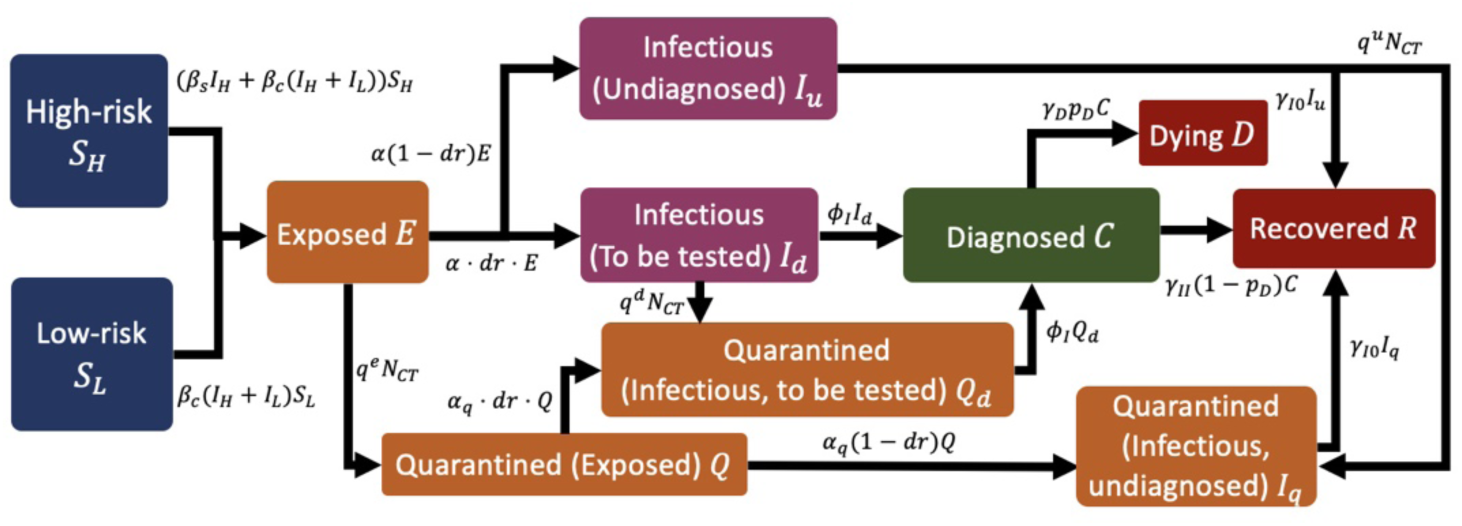
Model schematic. *I*_*H*_ = *I*_*uH*_ + *I*_*dH*_ + (1 − *c*_*iso*_)(*C*_*H*_ + *Q*_*dH*_ + *I*_*qH*_) and *I*_*L*_ = *I*_*uL*_ + *I*_*dL*_ + (1 − *c*_*iso*_)(*C*_*L*_ + *Q*_*dL*_ + *I*_*qL*_) refer to the effective infectious high-risk active and low-risk subpopulations, respectively, where *c*_*iso*_ is the reduction in capability of disease spreading due to quarantine or isolation and equals 1 in the absence of them. *N*_*CT*_ is the number of traced close contacts, whose value depends on contact tracing capacity, number of newly diagnosed cases and existing infected population. See Table S1–S2 for more details for these parameters.

To account for the possible disparity in transmission potential of mpox via community and additional sexual contacts and heterogeneity in contact patterns within the population, we segregated the affected population into two groups: high-risk (*H*) and low-risk (*L*) individuals. Compared to low-risk individuals (the general population), high-risk people were assumed to have an increased infection risk due to higher contact rates within this subpopulation, primarily driven by more frequent sexual contacts. Diagnostic rate and disease severity were not distinguished between these two groups.

Infections who underwent clinical testing would be diagnosed and reported, while those untested were assumed to have mild symptoms and would recover without medical assistance. Both groups were assumed the same infectious period, which was later varied in a sensitivity analysis (Figure S1–S2). We further stratified the diagnosed cases by severity. The majority (97%) were expected to have mild symptoms and be able to recover, while the remaining 3% were at high risk of death [2], with their survival status depending on the accessibility of medical resources.

Intervention measures built into the model include isolating diagnosed cases and quarantining the close contacts of confirmed cases. Both groups were presumed to have limited daily interactions with others. Please refer to section 2.3 and the Supplementary Information for more details.

### 2.2 Outbreak simulations for Asian cities

Utilising the established compartmental model, we simulated potential mpox outbreaks in 37 major municipals across Asia, each being either the capital or with the largest population size in its respective territory. We set the average number of secondary infections generated by an infector through community and additional sexual contacts in a fully susceptible population to be 1.02 (*R*_*c*_) and 1.62 (*R*_*s*_), respectively, implicitly presuming uniform baseline disease transmissibility across different territories. These values were derived from the estimated overall effective reproduction number and contributions of the two transmission routes during the 2024 mpox outbreak in South Kivu [16]. This extrapolation assumed that Clade Ib MPXV transmission in the local community in Asian cities would mirror the patterns observed in South Kivu except for the population-level immunity from smallpox vaccination or composition of high- and low-risk individuals. Using this combination of *R*_*s*_ and *R*_*c*_ and other pre-determined parameter values (Table S1–S2), we simulated the number of deaths and compared the model predictions with the confirmed death counts in South Kivu, the DRC [17] to validate this model at least within its original context (Figure S3–S4, Table S3–S4). Our simulated trajectory traced the index animal-to-human transmission event back to September 2023, which is consistent with findings in literature [1], substantiating the plausibility of the model and the corresponding parameter values used in projection.

Given the uncertainties surrounding travel volumes and interactions between travellers and local residents, we initiated the importation-triggered outbreaks by modelling the outcome of a one-time importation event, in which one or more susceptible individuals in the city were converted to exposed status. These individuals are henceforth referred to as *initial local infections*. We assumed no pre-existing exposed or active infections in the population prior to the importation. Neither did we account for cross protection from previous mpox outbreaks, due to the limited exposure to previous waves among individuals outside the GBMSM community and the few documented mpox cases in the region [5]. However, protection from smallpox vaccines were considered given their high efficacy and proven ability to prevent mpox infections, particularly among individuals aged over 45 years [18,19]. In our model, smallpox vaccines were treated as a leaky vaccine, where a reduced risk of infection was assumed among vaccinated individuals [20], while the alternative assumption of an all-or-nothing vaccine was evaluated in a sensitivity analysis (Figure S5–S6). We obtained the territory- and age-specific vaccination rates in 2024 using the model developed by Taube et al. [21], and aggregated the overall population immunity level using United Nations age distribution data [22] and a geospatial-invariant 80.7% vaccine effectiveness [21], based on which we scaled *R*_*s*_and *R*_*c*_ to obtain city-specific risks of infection.

Following the approach of Murayama et al. [16], we approximated the size of the high-risk subpopulation by considering the number of sex workers and their male clients, as they were more likely to engage in higher frequencies of sexual activity compared to the general population, thus more likely to contract the disease or infect others. Territory-specific sex worker statistics were collected from UNAIDS and the International Union of Sex Workers (IUSW) [23,24]. while the estimated proportions of clients among males aged 15–49 years were obtained from Carael et al. [25]. We further assumed that males comprised half of the population in each city and that all high-risk individuals fell in the age group of 15–49 years. Based on these assumptions, we calculated the proportions of sexually active individuals at high risk in each territory and their corresponding immunity level. To account for the potential bias in the territory-specific estimates due to limited data and potential temporal changes in the statistics [25], we additionally conducted a sensitivity analysis to explore how variations in the high-risk population could shape the outbreak dynamics in each city (Figure S7–S10).

We first simulated the outbreaks initiated by a single local infection resulting from a one-time importation event, modelling the scenarios in which the index infection occurred in either the low- or high-risk subpopulation. To assess the impact of importation size on disease transmission, we generated epidemic curves for the initial local infection sizes of one, three, and five. We set the probability of each initial local infection belonging to the high-risk group equal to the proportion of high-risk individuals within the overall population, assuming that initial local infections were all acquired through community contact. Nevertheless, we performed a sensitivity analysis to test this assumption, in which varying numbers of initial infections belonged to the high-risk group (Figure S11–S13). We did not assume any subsequent importation events or the implementation of NPIs (e.g., isolation or quarantine) in these scenarios.

### 2.3 Intervention effect projection

We evaluated two community-level NPIs: mandatory stay-at-home requirement for diagnosed cases (henceforth referred to as ‘isolation’) and quarantine of their close contacts captured by contact tracing (henceforth referred to as ‘quarantine’). These measures aimed at segregating the susceptible population from the detected infections and individuals exposed but not yet infectious, thereby reducing the risk of secondary infection.

Two intervention strategies, including

1) isolation alone, and
2) isolation combined with quarantine

were proposed and assessed under the scenario of three initial local infections due to a one-time importation event. For each strategy, four levels of quarantine or isolation effectiveness were assessed, in which the segregation was expected to reduce contacts between the susceptible and quarantined or isolated individuals by 25%, 50%, 75%, or 100%. The varying isolation effectiveness levels would allow for potential secondary infection of close contacts due to incomplete adherence, particularly in the case when community transmission be an established pathway of transmission [10]. Further details regarding these NPIs are elaborated in the Supplementary Information.

We simulated epidemic trajectories over a 5-year (i.e., 60-month) period for all the aforementioned scenarios, from which we derived statistics to reflect epidemic growth and outbreak size, including cumulative infections, cases, or potential deaths within one or five years following the initial local infection(s). Region-level trends were summarized as medians and interquartile ranges (IQR) of the 37 city-specific projections. All the analyses and visualisation were performed using the R software [26]. Data and analytical scripts are available at https://github.com/ShihuiJin/mpox_SEIR [27].

### 2.4 Patient and public involvement

### None

## 3. RESULTS

### 3.1 Susceptibility profiles of the 37 territories

The size of the susceptible population varied substantially across the 37 territories. Under the assumption of an 80.7% vaccine effectiveness, 34 territories had higher immunity levels due to smallpox vaccines compared to the DRC, with 27 having 75%–90% of their population remaining susceptible to MPXV. Territories in East Asia, including China, Hong Kong SAR, Japan, South Korea, and Taiwan, had the lowest percentages of susceptible population, at 60%–70%. Nevertheless, the proportion of high-risk susceptible individuals was relatively larger in theses territories, with at least 1.5% belonging to this subpopulation. Several Southeast Asian territories, such as Thailand, Cambodia, Laos, and Singapore, also had relatively high proportions of high-risk susceptible subpopulations, accounting for over 2% of the total population. This was particular in Thailand, where the proportion reached a high value of 2.9%. In contrast, the high-risk susceptible subpopulation was relatively small in many territories in Central and West Asia, hovering around or even below 1% (Figure 2).

**Figure 2.**
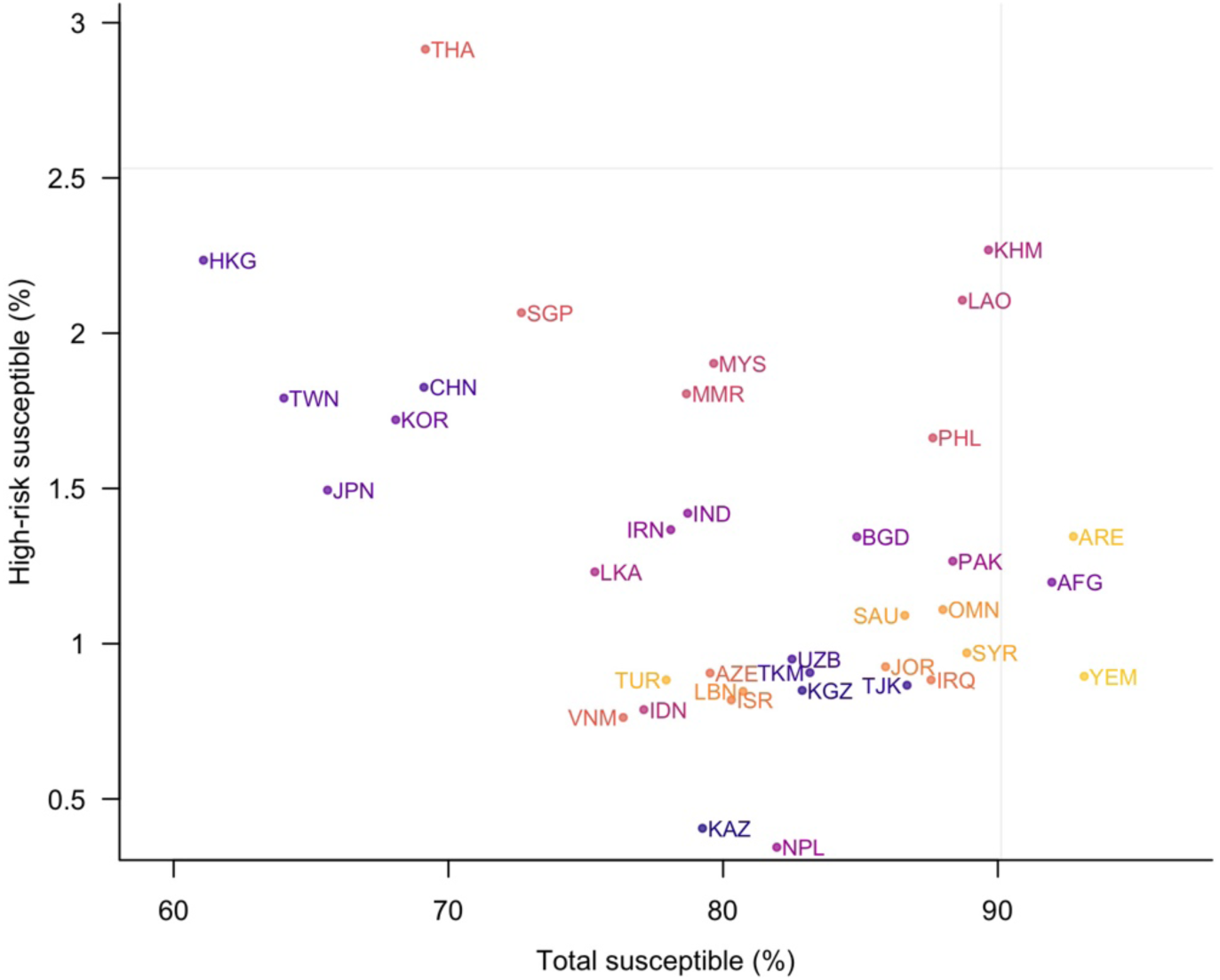
Proportion of total susceptible population (x-axis) and that of high-risk susceptible individuals (y-axis) in each territory. A vaccine effectiveness rate of 80.7% was assumed for smallpox vaccines. Territories within the same subregion (Central, East, Southeast, Southern, or West Asia) are represented by similar colours. The grey horizontal and vertical lines indicate the respective statistics in the DRC. Please refer to Table S2 for full names of the territories.

### 3.2 Outbreak scale under the scenario involving one initial local infection

When the index local infection with Clade Ib MPXV occurred in the low-risk group, Phnom Penh and Vientiane were identified as the two cities with highest risk of large-scale outbreaks, with projected case counts exceeding 50 within one year following the index local infection. A significantly larger outbreak size would be expected were the index local infection a high-risk individual, with over 200 cases expected in any of the 37 cities within the same timeframe. Four cities—Phnom Penh, Dubai, Vientiane, and Manila—were projected to experience outbreaks with over 1500 cases, while a few other cities in East and Southeast Asia, such as Taipei, Hong Kong, and Jakarta, would be relatively less affected (Figure 3). However, the ultimate scale of the outbreak by the end of the wave was minimally influenced whether the initial local infection occurred in the high- or low-risk group (Figure S14).

**Figure 3.**
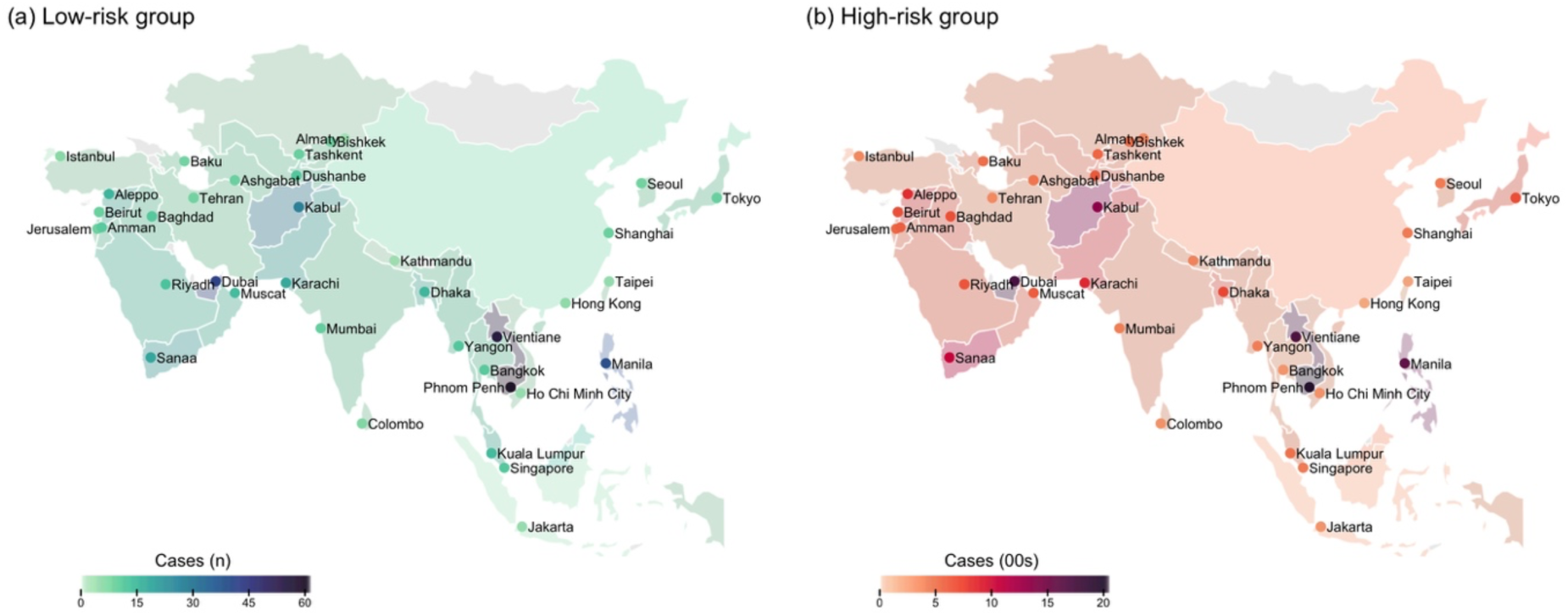
Outbreak sizes in one year. They are measured by total number of infections in one year following the initial local infection when the infection belonged to either the (a) low- or (b) high-risk group, across the 37 cities.

### 3.3 Impact of importation size on local transmission

The risk of experiencing large-scale outbreaks remained consistent across scenarios with varying initial local infection sizes, with number of infections, cases, or deaths in one year linearly correlated with the number of initial local infections. Specifically, in any city, an outbreak triggered by five initial local infections would result in approximately five times the number of infections within one year compared to an outbreak initiated by a single local infection (Figure 4).

**Figure 4.**
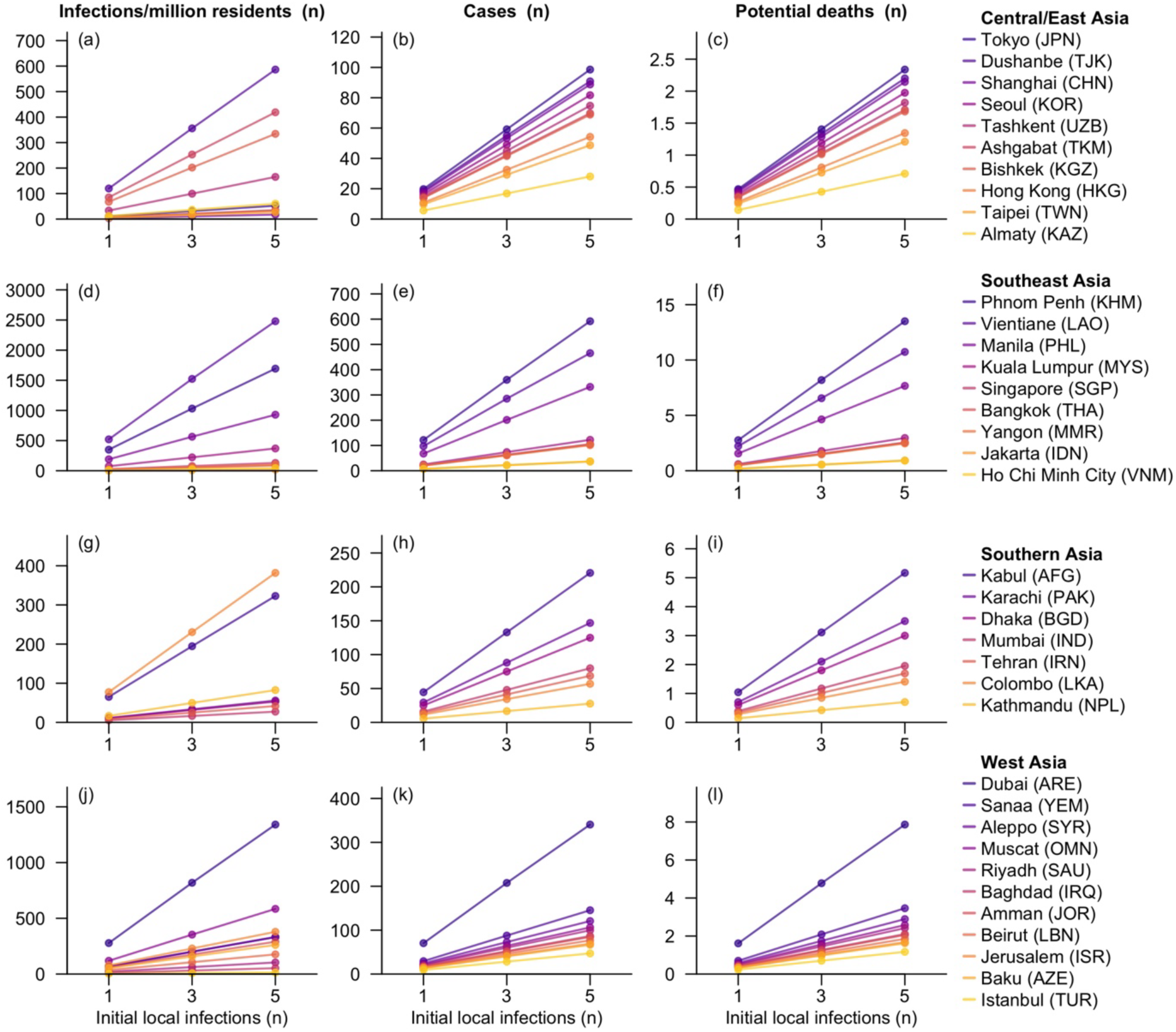
Summary statistics for simulated outbreaks in 37 Asian cities. The three statistics reported are: number of infections (Column 1), confirmed cases (Column 2), and individuals at risk of dying (Column 3) per million residents within one year following the initial local infections caused by a single importation event. The scenarios considered for each city include one, three, and five initial local infections. Cities within each subregion were ordered by potential death counts in the scenario with five initial local infections.

In three cities in Southeast and West Asia, including Vientiane, Phnom Penh, and Dubai, five initial local infections would lead to over 1000 infections per million residents within one year after the importation event that caused these infections. In contrast, significantly smaller proportions of the population would be affected in East Asian cities, with no more than 60 infections per million residents during the same period. Even under this hypothetical worst-case scenario, mortality would remain low in the first year, with no more than five infections per million residents at risk of dying was projected in 34 of the 37 Asian cities (Figure 4). However, it is worth noting that surges in case counts were not observed in countries until over one year after the initial local infections, and that the final outbreak size in one city by the end of the epidemic wave was marginally affected by the initial local infection sizes. Municipals with large populations or considerable number of high-risk individuals, such as Shanghai and Bangkok, were likely to experience high case counts during the wave, even if the outbreak appeared relatively modest in the first year (Figure S15).

Note that the projected epidemic trajectories might vary substantially if alternative high-risk population sizes were assumed instead of the values reported in literature (Figure S8–S10). Furthermore, the sensitivity analyses also show that the outbreak size in one year following the initial local infections would be affected by the number of initial infections in the high-risk population, where a higher proportion of infections falling in this group would substantially increase the outbreak scale (Figure S11–S13). Modelling the smallpox vaccine as an all-or-nothing mechanism, rather than a leaky vaccine as assumed in the main analysis, would also result in more infections over the first year (Figure S5–S6). However, a potentially shortened infectious period among asymptomatic or mild infections could delay outbreaks and reduce the scale across the Asian cities (Figure S1–S2). Please refer to the Supplementary Information for further details on the simulated outbreaks under these alternative scenarios.

### 3.4 Effectiveness of NPIs on outbreak control

Both isolation and quarantine would contribute to significant reductions in confirmed cases for the 37 cities. In the scenario with three initial local infections, compared to the baseline without NPIs, isolation alone would bring a 43.8% (IQR: 42.8%–44.7%), 67.8% (IQR: 66.5%–68.9%), 80.8% (IQR: 79.5%–82.0%), and 88.0% (IQR: 86.8%– 89.1%) reduction to the cumulative cases within one year following the initial local infection events, when it decreased the interpersonal contacts by 25%, 50%, 75%, and 100%, respectively. Adding quarantine would further lower the cumulative cases by 22.0 (IQR: 21.8–22.1), 19.9 (IQR: 19.6–20.0), 13.5 (IQR: 13.1–13.7), and 8.4 (IQR: 8.0–8.8) percentage points for the four levels of isolation effectiveness (Figure 5). Nevertheless, the impact of isolation substantially diminished when the assessment window was extended to five years, while a significant increase was observed in the effectiveness of quarantine, potentially due to the delayed outbreak surge caused by interventions (Figure S16–S17, Table S5).

**Figure 5.**
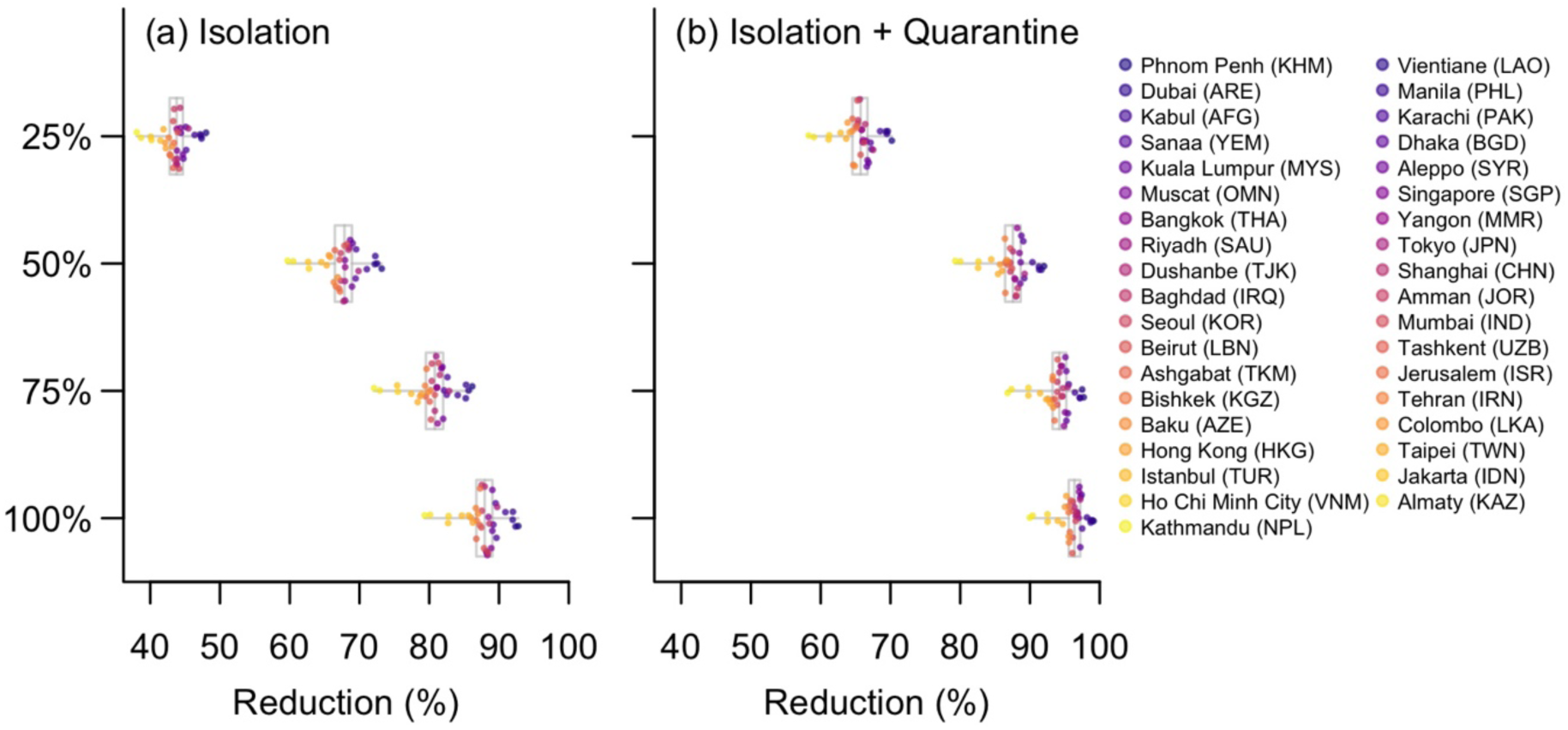
Projected reduction in confirmed cases in one year following the initial three local infections due to (a) isolation or (b) isolation combined with quarantine. The reduction proportions were calculated for each level of isolation effectiveness (25%, 50%, 75%, and 100% reduction in interpersonal contacts). compared to the baseline scenario with no NPIs and three initial local infections per city. Grey box plots display the median, interquartile ranges, and ranges of reductions across the 37 cities. In the legend bar on the right, these cities were ordered by confirmed case counts in one year in the scenario without NPIs, with a darker colour indicating a greater decrease.

The impacts of NPIs also varied greatly across the 37 cities. Cities experiencing larger outbreaks within the first year tended to benefit more from NPIs. For example, in Phnom Penh, isolation alone could decrease 48.0%, 73.2%, 86.2%, and 92.8% of confirmed cases at the four effectiveness levels, respectively. Conversely, in cities like Kathmandu and Almaty, where the fewer than 20 cases were expected in the first year, reductions were lower by at least nine percentage points at each effectiveness level (Figure 5, Figure S18). Nevertheless, this strong association between intervention effectiveness and local outbreak size was less evident when the assessment period was extended to five years (Figure S16, S19).

## 4. DISCUSSION

In this study, we simulated localized mpox outbreaks triggered by a single importation event to project the potential spread of Clade Ib MPXV in 37 Asian cities with varying population sizes, levels of immunity from smallpox vaccination, and proportions of sexually active population at higher transmission risk. In the scenario involving three initial local infections, an outbreak of up to 340 cases per million residents was anticipated within one year in the absence of interventions, while the implementation of potent isolation and quarantine measures could lower the incidence to fewer than four per million.

Our simulation results revealed positive correlations between outbreak sizes and both the proportion of sexually active individuals at high transmission risk and the level of immunity due to smallpox vaccination in each city. While the large-scale outbreaks in Dubai was mainly attributed to the low vaccine-induced (7%), the substantial number of infections projected for Thailand, where smallpox vaccines were estimated to protect over 30% of the population from MPXV infections, was primarily the result of the high proportion of high-risk individuals in the community [21]. Specifically, over 10% of males aged 15–49 years were documented to engage with sex workers [25], leading to an estimated 2.9% of the population being unvaccinated and categorised as high-risk in our study (Figure 2). This finding is further supported by the results of the sensitivity analysis, where Bangkok was predicted to be among cities with the lowest outbreak risks under the assumption of a uniform high-risk proportion among individuals aged 15–49 years across different cities (Figure S7–S10).

Both NPIs assessed in our analysis—case isolation, and quarantining of close contacts—demonstrated promising potential in mitigating the outbreaks across all Asian cities. While median reduction in the total case counts over one year was expected to approach roughly 90% in the ideal scenario where isolation was 100% effective in preventing contacts between susceptible and isolated infectious individuals, the rate of case reduction would diminish per 25% increment in isolation effectiveness. The actual effectiveness would depend on stringency of isolation policies, the isolation facilities required (e.g., home or designated healthcare centres), and compliance of affected individuals, which may vary between countries. Nevertheless, the moderate simulated outbreak sizes in most cities suggest the feasibility of less restrictive isolation measures, such as home isolation for confirmed cases, since even with 50% isolation effectiveness, our model projected fewer than 100 confirmed cases per million residents in any Asian city in one year following three initial local infections (Figure S17–S18). Furthermore, given the significantly longer infectious period of mpox compared to COVID-19 [15], extended isolation in healthcare facilities could easily strain pre-existing infrastructure with a surge in case counts.

The added benefits of quarantining close contacts, beyond isolation alone, was consistently projected to slowing the disease spread, contributing to an additional reduction of at least five percentage points in the cases counts over one year. This aligns with its extensive use during the COVID-19 pandemic [28,29]. However, delays in contact tracing may affect quarantine effectiveness. In our analysis, we assumed a uniform three-day interval between diagnosis of index cases and the identification of their close contacts [30], but longer delays in contact tracing could result in additional secondary infections caused by the infectious close contacts. It is also important to note that effective contact tracing would require substantial human resources for identifying close contacts, which may not always be readily available. As such, isolation is recommended as the primary intervention strategy for containing all potential outbreaks, with contact tracing and quarantining serving as supplementary measures in scenarios involving relatively large outbreak scales.

It is worth highlighting that our estimates of outbreak sizes and disease burden were based on simulated epidemic trajectories initiated by a one-time importation, whose outcomes were modelled through a fixed size of initial local infections in the fully susceptible local population for each city. It should be noted that our simulations assumed these initial infections were contracted through community contact. This assumption may underestimate the number of infections over one year should the initial local transmission occur through sexual contact within the high-risk group (Figure S11–S13). Moreover, we did not account for recurring importation events due to the observation of single imported cases in individual affected countries outside the African continent by September 2024. Challenges also exist in quantifying the infectivity of incoming infectious travellers on local residents, as MPXV is mainly transmitted through close contacts [10] and contact patterns between travellers and the local population might differ significantly from those within the local community. Nevertheless, given the high disease transmissibility, especially through sexual transmission, the role of importation would likely be minimal in influencing the epidemic curve once the virus begins to circulate within the city.

In this study, we employed a deterministic model to project the average outcome of disease transmission across diverse scenarios. The model’s lack of stochasticity limits the its capability to capture the uncertainties in the epidemic trajectories. Particularly, in low-incidence scenarios with fewer than five infections over an extended period, stochastic effects in reality could lead to the extinction of infections, while a deterministic model allows for fractional infections and prevents the transmission from dying out as long as the effective reproduction number is at least one. This poses challenges in quantifying the probability of extinction and interpreting predictions during the early stages of an outbreak. Therefore, we alternatively focused on infection sizes over an extended simulation period, during which the outbreak sizes were generally sufficiently large. In this context, the consistent, predictable outcomes generated by the deterministic model offer a more intuitive quantification of the potential outbreak scales in the investigated cities, supporting comparisons across settings and facilitating informed decision-making.

A further simplification of real-world scenarios in our study lies in the modelling of vaccination effectiveness. We assumed a leaky vaccine model for historical smallpox vaccines, representing their impact as a population-level reduction in disease transmissibility. Compared to an all-or-nothing assumption, this assumption led to slower outbreak progression but a larger final outbreak size (Figure S5–S6) [31]. Meanwhile, we did not account for the reducing population immunity due to aging over the long simulation window, which may have caused an underestimation of the final outbreak sizes.

In addition, it should be noted that our approximation of high-risk population sizes was based on reported numbers of sex workers and their clients [23–25]. Although consistent data sources were utilized for most territories and the same calculation method was applied to enhance the comparability across territories, relevant statistics were unavailable for certain territories, especially those in the Middle East. Consequently we extrapolated the figures using subregional average proportions, which were lower than the reported values for other parts of Asia.

Possibilities also exist that the number of individuals engaging with sex-workers was under-reported in surveys due to strict cultural norm in some Asian territories and the stigma associated with acknowledging commercial sex [25,32]. Both factors could lead to an underestimation of the high-risk population sizes and hence an underestimation of outbreak size, as sexual transmission was a key driver of disease spread in our simulations.

Another major limitation of our study pertains to the large uncertainty surrounding transmission-related model parameters. The co-circulation of multiple MPXV clades in Africa [33], along with a lack of comprehensive sequencing data, complicates efforts to pinpoint the transmissibility of MPVX Clade Ib. The disparities in transmission dynamics across regions with reported Clade Ib MPXV infections introduced additional unsureness regarding the contribution of sexual transmission to the overall outbreak [16]. In this study, we stratified the population according to their sexual activity levels and based the transmissibility estimates for these two groups on disease surveillance data from South Kivu, assuming that transmission in the province was predominately driven by Clade Ib MPXV. These parameter estimates were derived using a next generation matrix approach, accounting for the impacts of heterogenous sexual networks and the age-specific contact patterns [16], but our population-level model did not fully capture the detailed network structure. Since the cases identified outside the African continent primarily consisted of importations and data on local transmission remained scarce by September 2024, we extrapolated these estimates from the outbreak data in the DRC to the Asian urban setting without validating against context-specific observations. We characterized the heterogeneity in effective reproduction number across individual cities using territory-specific smallpox immunity profiles and high-risk population sizes. However, several other driving factors of the transmission potential were not included due to the limited available data and challenges in quantifying their exact impacts on shaping the transmissibility of Clade Ib MPXV. Particularly, geospatial differences are likely to exist in the number and frequency of close contacts across the 37 Asian cities, including both sexual interactions and physical contacts within households stemming from discrepancies in family sizes, demographical structures, living habits, cultural norms, and other social and economic factors. This limits the generalisability of our results. Given the relatively smaller household sizes in many Asian cities compared to those in the DRC [34], the number of infections might have been overestimated. The shortened viral shedding duration among asymptomatic or mild infections [35], coupled with their potentially reduced frequencies of contacts owing to self-isolation, may also contribute to an overestimation of their infectivity and hence an overestimation of outbreak sizes (Figure S1–S2). Furthermore, the reliance on surveillance data sourcing from Africa, where healthcare and monitoring systems may face constraints, might have led to an underestimation of infection sizes and an overestimation of disease severity. Consequently, the projected potential death counts based on African data in our study may far exceed reality in many Asian cities. With the availability of more case data and epidemiological parameters, our model may require updating.

Despite these limitations, our simulations suggest that while mpox outbreaks triggered by importation could lead to substantial morbidity and mortality in an Asian city with large populations of sexually active individuals at risk, they would still remain controllable provided adequate preparation and a timely response from decision-makers, underscoring the significance of robust surveillance systems. In addition, the transmission model proposed in this study enables the quantification of impacts of containment measures widely applied during the COVID-19 pandemic on mpox outbreaks, an area much less explored compared to the virus’s transmission patterns [1,36]. Provided proper implementation, the NPIs evaluated in this study have the potential to suppress outbreak sizes and curb disease spread by up to 99%, offering evidence for adopting them to effectively manage new outbreaks with potentially high community transmissibility. Based on these findings, there exists an urgent need for strong surveillance systems, efficient contact tracing, quarantining of close contacts, and institutional isolation of cases.

## Data Availability

All data produced in the present study are available upon reasonable request to the authors.

## Author contributions

SJ, GG, and BLD conceived and designed the study. SJ implemented the statistical analysis and created the figures and tables. SJ wrote the original draft of the manuscript. GG, AE, KP, RKJT, JTL, KE, and BLD reviewed and edited the manuscript. BLD is the guarantor.

## Data and code availability

Supplementary data and analytical codes utilized for this study are available at https://github.com/ShihuiJin/mpox_SEIR.

## Ethics

No Ethics Approval is needed for this study.

## Protocol

No protocol is prepared for this study.

## Registration

The study is not registered.

