## Supplementary Information for "Modelling the potential spread of Clade Ib MPXV in Asian cities"

### Additional Explanations for the Model and Proposed Scenarios

**Table S1. Name, definition, and derivation approach (where appliable) of the model parameters.**

| **Parameter** | **Definition** | **Value/Source** |
| --- | --- | --- |
| *Population-related* |  |  |
| $N$ | Overall population size | Table S2; From literature [1] |
| $p_{H}$ | Proportion of high-risk individuals in the general population | Table S2; From literature [2–4] |
| $p_{L}$ | Proportion of low-risk individuals in the general population | $1-p_{H}$ |
| $N_{H}$ | High-risk population size | $p_{H}N$ |
| $N_{L}$ | Low-risk population size | $p_{L}N$ |
| *Transmission-related* |  |  |
| $R_{s}$ | Average number of secondary infections generated by an infector (from the high-risk group) through sexual contacts in addition to the general community contacts in a population with no previous exposure to the disease | 1.62; From literature [5], in which the effective reproduction number for additional sexual transmission was estimated to be 1.51^#^, with 93.4% of the population remaining susceptible |
| $R_{c}$ | Average number of secondary infections generated by a typical infector in the general community with no previous exposure to the disease | 1.02; From literature [5], in which the effective reproduction number for transmission in the general community was estimated to be 0.92^#^, with 90.1% of the population remaining susceptible |
| $s_{H}$ | Susceptibility to Clade Ib monkeypox virus (MPXV) among high-risk individuals | Table S2; From literature [6] |
| $s$ | Population-level susceptibility to Clade Ib MPXV | Table S2; From literature [6] |
| $\beta_{s}$ | Rate of disease transmission through sexual contacts in addition to the general community contacts | $s_{H}\gamma_{I}R_{s}/N_{H}$ |
| $\beta_{c}$ | Rate of disease transmission through community contacts | $s\gamma_{I}R_{c}/N$ |
| $c_{iso}$ | Proportion of reduction in the number of secondary infections due to quarantine or isolation | 0 without intervention (default), and 25%, 50%, 75%, or 100% with intervention; Assumed |
| $\alpha$ | Rate of latent individuals becoming infectious | Inverse of average duration from initial exposure to symptom onset (7d); From literature [7] |
| $\gamma_{I0}$ | Recovery rate for the undiagnosed infectious | Inverse of average recovery period (21d); From literature [8] |
| $\gamma_{II}$ | Recovery rate for the diagnosed cases | Inverse of the average duration from diagnosis to recovery/ loss of infectiousness (15d); From literature [8] |
| $\gamma_{D}$ | Death rate | Inverse of the average duration from symptom onset to death (15d); From literature [9], assuming hospitalized patients who did not recover within the average period would have server symptoms and of higher risks of dying |
| *Severity-related* |  |  |
| $p_{D}$ | Case-fatality risk | 3.0%; From literature [10] |
| *Surveillance-related* |  |  |
| $dr$ | Diagnostic rate | 25%; From literature [11] |
| $\phi_{I}$ | Rate of diagnosis | Inverse of average duration from symptom onset to diagnosis (6d); From literature [12] |
| $ct_{max}$ | Maximum number of new cases to perform contact tracing per day | 0 without intervention (default) and 30 with intervention; Assumed |
| $ct_{connect}$ | Average number of close contacts traced per new case | 1; Assumed based on the average number of secondary infections generated by an infectious individual |
| $N_{CT}$ | Number of traced, infected close contacts | Capped at the minimum of exposed population and $(ct_{max}\cdot ct_{connect})$ |
| $q^{e}$ | Proportion of traced, latent close contacts | Proportion of exposed individuals among all infections not yet diagnosed or quarantined |
| $q^{d}$ | Proportion of traced, infectious and to be diagnosed close contacts | Proportion of traced, infectious and to be diagnosed individuals among all infections not yet diagnosed or quarantined |
| $q^{u}$ | Proportion of traced, infectious close contacts who would remain undiagnosed until recovery | Proportion of traced, infectious individuals who would remain undiagnosed until recovery among all infections not yet diagnosed or quarantined |
| $\alpha_{q}$ | Rate of latent individuals becoming infectious among individuals in quarantine | Inverse of average duration from quarantine initiation to symptom onset (4d), adjusted by delay in contact tracing (3d); From literature [13] |

^#^ Having been adjusted to account for population-level immunity from smallpox vaccination. We assumed a leaky vaccine [14] and used the estimated age-specific smallpox vaccine coverage in the Democratic Republic of the Congo (DRC), the United Nations age distribution data, and an 80.7% vaccine effectiveness [6,15]. This led to our estimation that 90% of the general population and 93% of the high-risk individuals (aged 15–49) remained susceptible to mpox at the start of an outbreak.

**Table S2. City-specific information.** This includes city name, corresponding ISO3 code and territory name, population size ($N$), population-level susceptibility to Clade Ib MPXV ($s$), proportion of high-risk individuals in the general population ($p_{H}$), and susceptibility to Clade Ib MPXV among the high-risk individuals ($s_{H}$). The population-level susceptibility was derived based on the estimated age-specific smallpox vaccine coverage ($p_{vac}$) and demographics in individual territories, under the assumption of a leaky vaccine (i.e., calculated as $\left( 1-ve\cdot p_{vac} \right)\cdot100\%,$ where $ve=80.7\%$ is the assumed smallpox vaccine effectiveness).

| **City** | **ISO** | **Territory** | **N (million)** | $\boldsymbol{s}$ **(%)** | $\boldsymbol{p}_{\boldsymbol{H}}$ **(%)** | $\boldsymbol{s}_{\boldsymbol{H}}$ **(%)** |
| --- | --- | --- | --- | --- | --- | --- |
| Aleppo | SYR | Syrian Arab Republic | 1.60 | 88.9 | 1.0 | 94.1 |
| Almaty | KAZ | Kazakhstan | 2.00 | 79.2 | 0.4 | 90.9 |
| Amman | JOR | Jordan | 1.28 | 85.9 | 1.0 | 92.2 |
| Ashgabat | TKM | Turkmenistan | 0.73 | 83.2 | 1.0 | 91.5 |
| Baghdad | IRQ | Iraq | 7.22 | 87.6 | 1.0 | 92.2 |
| Baku | AZE | Azerbaijan | 1.12 | 79.5 | 1.0 | 92.2 |
| Bangkok | THA | Thailand | 5.10 | 69.2 | 3.3 | 87.5 |
| Beirut | LBN | Lebanon | 1.92 | 80.7 | 0.9 | 93.7 |
| Bishkek | KGZ | Kyrgyzstan | 0.90 | 82.9 | 0.9 | 92.1 |
| Colombo | LKA | Sri Lanka | 0.65 | 75.3 | 1.4 | 89.1 |
| Dhaka | BGD | Bangladesh | 10.4 | 84.9 | 1.5 | 92.4 |
| Dubai | ARE | United Arab Emirates | 1.14 | 92.8 | 1.4 | 99.5 |
| Dushanbe | TJK | Tajikistan | 0.68 | 86.7 | 0.9 | 93.4 |
| Ho Chi Minh City | VNM | Viet Nam | 3.47 | 76.4 | 0.9 | 88.8 |
| Hong Kong | HKG | Hong Kong SAR | 7.01 | 61.1 | 2.6 | 86.8 |
| Istanbul | TUR | Türkiye | 14.8 | 77.9 | 1.0 | 89.5 |
| Jakarta | IDN | Indonesia | 8.54 | 77.1 | 0.9 | 88.4 |
| Jerusalem | ISR | Israel | 0.80 | 80.3 | 0.9 | 93.3 |
| Kabul | AFG | Afghanistan | 3.04 | 92.0 | 1.2 | 96.7 |
| Karachi | PAK | Pakistan | 11.6 | 88.4 | 1.4 | 93.4 |
| Kathmandu | NPL | Nepal | 1.44 | 82.0 | 0.4 | 91.2 |
| Kuala Lumpur | MYS | Malaysia | 1.45 | 79.7 | 2.1 | 90.4 |
| Manila | PHL | Philippines | 1.60 | 87.6 | 1.7 | 98.5 |
| Mumbai | IND | India | 12.7 | 78.7 | 1.6 | 89.7 |
| Muscat | OMN | Oman | 0.80 | 88.0 | 1.2 | 91.9 |
| Phnom Penh | KHM | Cambodia | 1.57 | 89.7 | 2.3 | 99.7 |
| Riyadh | SAU | Saudi Arabia | 4.21 | 86.6 | 1.2 | 91.8 |
| Sanaa | YEM | Yemen | 1.94 | 93.1 | 0.9 | 95.2 |
| Seoul | KOR | Republic of Korea | 10.4 | 68.1 | 1.9 | 91.0 |
| Shanghai | CHN | China | 22.3 | 69.1 | 2.0 | 90.9 |
| Singapore | SGP | Singapore | 3.55 | 72.7 | 2.3 | 90.2 |
| Taipei | TWN | Taiwan | 7.87 | 64.0 | 2.1 | 87.2 |
| Tashkent | UZB | Uzbekistan | 1.98 | 82.5 | 1.0 | 91.8 |
| Tehran | IRN | Iran (Islamic Republic of) | 7.15 | 78.1 | 1.5 | 88.8 |
| Tokyo | JPN | Japan | 8.34 | 65.6 | 1.6 | 94.5 |
| Vientiane | LAO | Lao People's Democratic Republic | 0.84 | 88.7 | 2.1 | 98.9 |
| Yangon | MMR | Myanmar | 4.48 | 78.7 | 2.0 | 89.5 |

**Rationale for splitting the population into high- and low-risk groups**

In our model, residents in a city were divided into high- and low-risk subpopulations. High-risk individuals were assumed more likely to transmit or contract mpox due to a greater frequency of sexual contacts compared with their low-risk counterparts. This categorization was informed by the definition of $R_{s}$ and $R_{c}$ sourced from the literature [5]. $R_{c}$ quantified the disease transmissibility in the general community, which was calculated using a survey-based social contact matrix that includes both sexual and non-sexual contacts. In contrast, $R_{s}$ represented the additional transmission potential attributed to frequent sexual contacts within the high-risk population.

**Transition between different compartments of the model**

Compartments in our model encompass Susceptible ($S_{H}$ for high-risk group and $S_{L}$ for low-risk group), Exposed ($E$), Quarantined ($Q)$, Infectious ($I_{u}$ for the undiagnosed, $I_{d}$ for those who had been tested but yet to be confirmed, and $C$for the diagnosed), Recovered ($R$), and Dead ($D$). In addition, compartments making up of the infectious population, including $I_{u}$, $I_{d}$, $C$, $Q_{d}$, and $I_{q}$ were further stratified into the high-risk and low-risk groups, which were denoted by subscripts $H$ and $L$, respectively. From these groups, we derived the number of infectious sexually active or inactive individuals, as $I_{H}=I_{uH}+I_{dH}+\left( 1-c_{iso} \right){(C}_{H}+Q_{dL}+I_{qH})$ and $I_{L}=I_{uL}+I_{dL}+\left( 1-c_{iso} \right)(C_{L}+Q_{dL}+I_{qL})$, respectively. $N_{CT}$ represents the number of traced close contacts in the scenario where quarantine was implemented. These close contacts were could be exposed but not yet infectious, infectious and to be diagnosed, or undiagnosed infectious individuals, accounting for $q^{e}$, $q^{d}$, and $q^{u}$ of the total $N_{CT}$, respectively (details in the subsequent section **Explanations for individual non-pharmaceutical interventions (NPIs)**). The transitions between compartments for local transmission are described as below. For simplicity, $X$ is used as a placeholder for $H$ or $L$ when both groups follow the same transition procedure between neighbouring states. Compartment labels without subscripts denote the total of all possible categories.

$$\frac{dS_{H}}{dt}=-\left( \beta_{S}I_{H}+\beta_{C}\left( I_{H}+I_{L} \right) \right) S_{H}$$

$$\frac{dS_{L}}{dt}= -\beta_{C}\left( I_{H}+I_{L} \right)S_{L}$$

$$\frac{dE_{H}}{dt}=\left( \beta_{S}I_{H}+\beta_{C}\left( I_{H}+I_{L} \right) \right) S_{H} -p_{H}{q^{e}N}_{CT}-\alpha E_{H}$$

$$\frac{dE_{L}}{dt}=\beta_{C}\left( I_{H}+I_{L} \right)S_{L} -p_{L}{q^{e}N}_{CT}-\alpha E_{L}$$

$$\frac{dQ_{X}}{dt}={p_{X}q_{X}^{e}N}_{CT}-\alpha_{q}Q_{X}$$

$$\frac{dI_{uX}}{dt}=\alpha E_{X}\cdot\left( 1-dr \right)-p_{X}q^{u}N_{CT}- \gamma_{I0}I_{uX}$$

$$\frac{dI_{dX}}{dt}=\alpha E_{X}\cdot dr-p_{X}q^{d}N_{CT}- \phi_{I}I_{dX}$$

$$\frac{dQ_{dX}}{dt}={p_{X}q_{X}^{d}N}_{CT}+\alpha_{q}Q_{X}\cdot dr- \phi_{I}Q_{dX}$$

$$\frac{dC_{X}}{dt}= \phi_{I}(I_{dX}+Q_{dX})-\gamma_{II}\left( 1-p_{D} \right)C_{X}-\gamma_{D}p_{D}C_{X}$$

$$\frac{dI_{qX}}{dt}={p_{X}q_{X}^{u}N}_{CT}+\alpha_{q}Q_{X}\cdot\left( 1-dr \right)- \gamma_{I0}I_{qX}$$

$$\frac{dR}{dt}= \gamma_{I0}I_{u}+\gamma_{I0}I_{q}+ \gamma_{II}\left( 1-p_{D} \right)C$$

$$\frac{dD}{dt}= \gamma_{D} p_{D}C$$

Importation was modelled by quantifying its impact on local population, i.e., through the reduction of the susceptible population and increase in the exposed population. We simulated the average effect of importation, assigning a fixed proportion of resulted local new infections to the high-risk group and the rest to the low-risk group. This proportion corresponds to the ratio of high-risk individuals in the susceptible population at the beginning of the simulation. Epidemic curves for the extreme scenarios, in which the index infection occurred in either high- or low- risk population, were also generated for the Asian cities when there is only one local infection resulted from importation events in the main analysis.

**Modelling pre-existing immunity from smallpox vaccination**

Let $p_{vac}$ denote the coverage of smallpox vaccines in a population, population-level susceptibility was calculated as

$$s=\left( 1-ve\cdot p_{vac} \right)\cdot100\%,$$

where $ve=80.7\%$ is the effectiveness of smallpox vaccines [6]. The impact of vaccination on transmission was modelled as a reduced risk of infection rather than complete immunity for a subset of the population. Consequently, the effective reproduction number for the population was the basic reproduction number scaled by $s$ (i.e., $sR_{0}$). A sensitivity analysis was performed to compare the impacts of these two approaches to model vaccination effects.

**Explanations for individual non-pharmaceutical interventions (NPIs)**

*Isolation*

All the diagnosed infections were assumed to start isolation immediately after diagnosis or being confirmed as a mpox case. When isolation occurred at home, there was a risk of household transmission and non-compliance. Consequently, we considered scenarios in which transmission risk was not eliminated. Specifically, we assumed the reduction in the average number of secondary infections generated by isolated infections to be 25%, 50%, 75%, and 100%. In scenarios without this intervention, the relevant parameter, $c_{iso}$, was set to 0.

*Contact tracing and quarantine*

The number of traced, infected close contacts ($N_{CT}$) on day $t$ was determined by the following factors: the number of newly diagnosed cases from three days prior ($\Delta D_{t-3}$), individuals currently exposed but not yet infectious ($E_{t-1}$), and infectious but not yet diagnosed individuals ($I_{d,t-1}$ for those to be diagnosed and $I_{u,t-1}$ for those who would remain undiagnosed until recovery), contact tracing capacity ($ct_{max}$), as well as the number of close contacts per case ($ct_{connect}$), i.e.,

$$\min(E_{t-1}+I_{d,t-1}+I_{u,t-1},\min(\Delta D_{t-3}, ct_{max})\cdot ct_{connect}).$$

The allocation of traced, infected close contacts between the high- and low-risk groups was determined by the number of infected but not yet diagnosed or quarantined individuals in each group, while the time-dependent parameters $q^{e}$, $q^{d}$, and $q^{u}$ were calculated based on the sizes of $E_{t-1}$, $I_{d,t-1}$, and $I_{u,t-1}$for the respective groups.

Considering the potential constraints on manpower for contact tracing, we set$ct_{max}=30$ as the spatio-temporally invariant maximum number of cases which could undergo manual contact tracing per day due to the limited resources available for this effort, and $ct_{connect}=1$ as the average number of unique traced close contacts exposed to Clade Ib MPXV per case, based on our assumed average number of secondary infections generated by each infectious individuals. A three days’ delay was assumed between the initiation of contact tracing and implementation of quarantine [13]. We assumed all the traced contacts were either exposed or infectious, with the proportion of infectious individuals matching that for the untraced population. We did not consider mandatory testing for individuals in quarantine, thereby applying the same diagnostic rate and risks of developing severe symptoms or dying to both the traced close contacts and the unquarantined local infections. In scenarios without this intervention, $N_{CT}=ct_{max}=0$.

### Sensitivity analysis: Impact of a reduced infectious period for undiagnosed cases

A large proportion of undiagnosed infections were asymptomatic or exhibited mild symptoms. Considering the potentially shorter shedding period associated with such infections [21], a sensitivity analysis was conducted to evaluate its impact on the projected epidemic trajectories. Specifically, the infectious period for undiagnosed cases was reduced to 14 days [22], compared to the 21-day infectious period assumed for diagnosed mild cases in our main analysis. This resulted in delayed outbreaks with significantly smaller outbreak sizes over the five-year period (Table S1–S2).

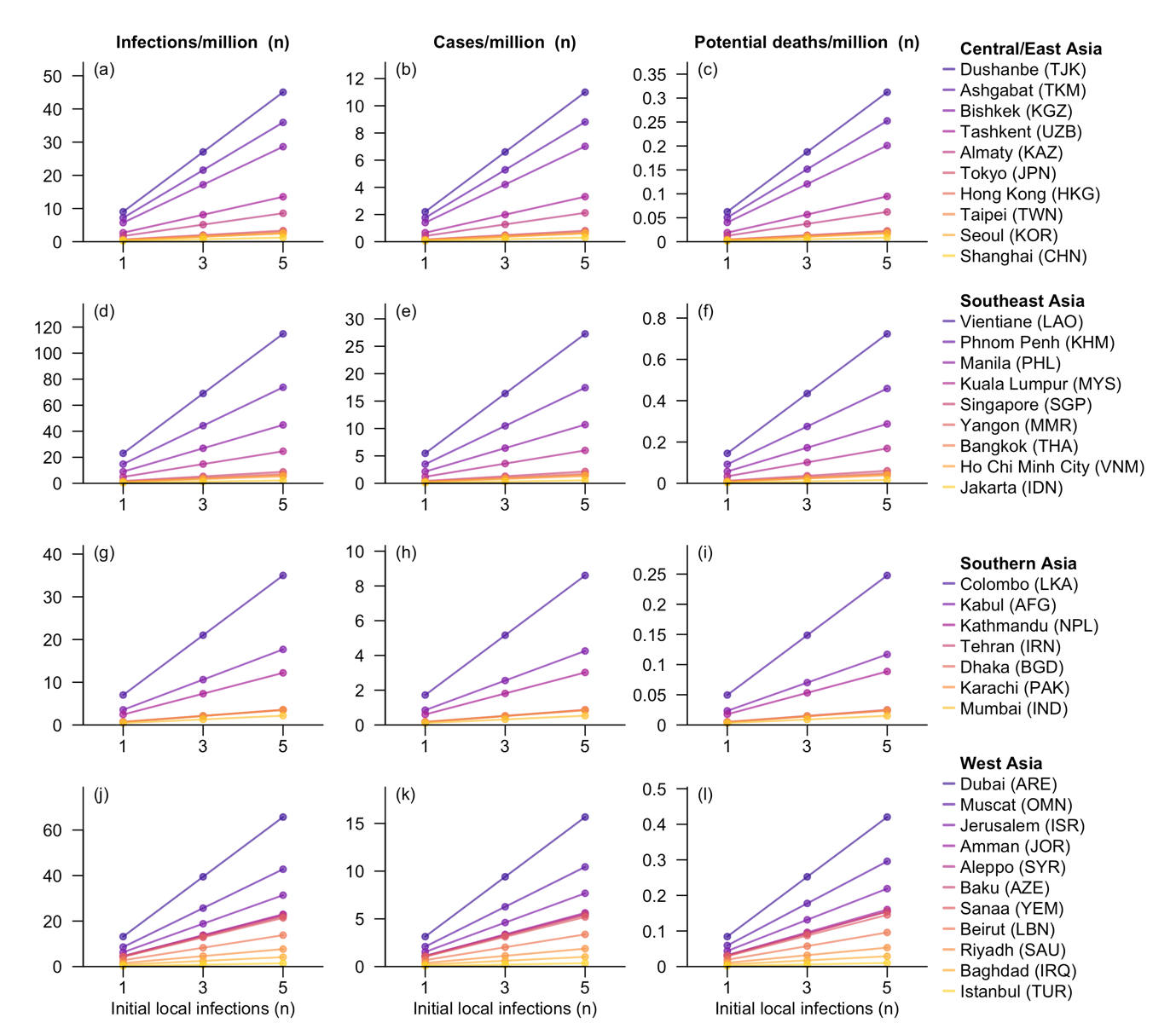
**Figure S1. Summary statistics for outbreaks in 37 Asian cities, assuming a 14-day infectious period for undiagnosed infections.** The three statistics reported are: number of infections (Column 1), confirmed cases (Column 2), and individuals at risk of dying (Column 3) per million residents within one year following the initial local infections caused by a single importation event. The scenarios considered for each city include one, three, and five initial local infections. Cities within each subregion were ordered by potential death counts in the scenario with five initial local infections.

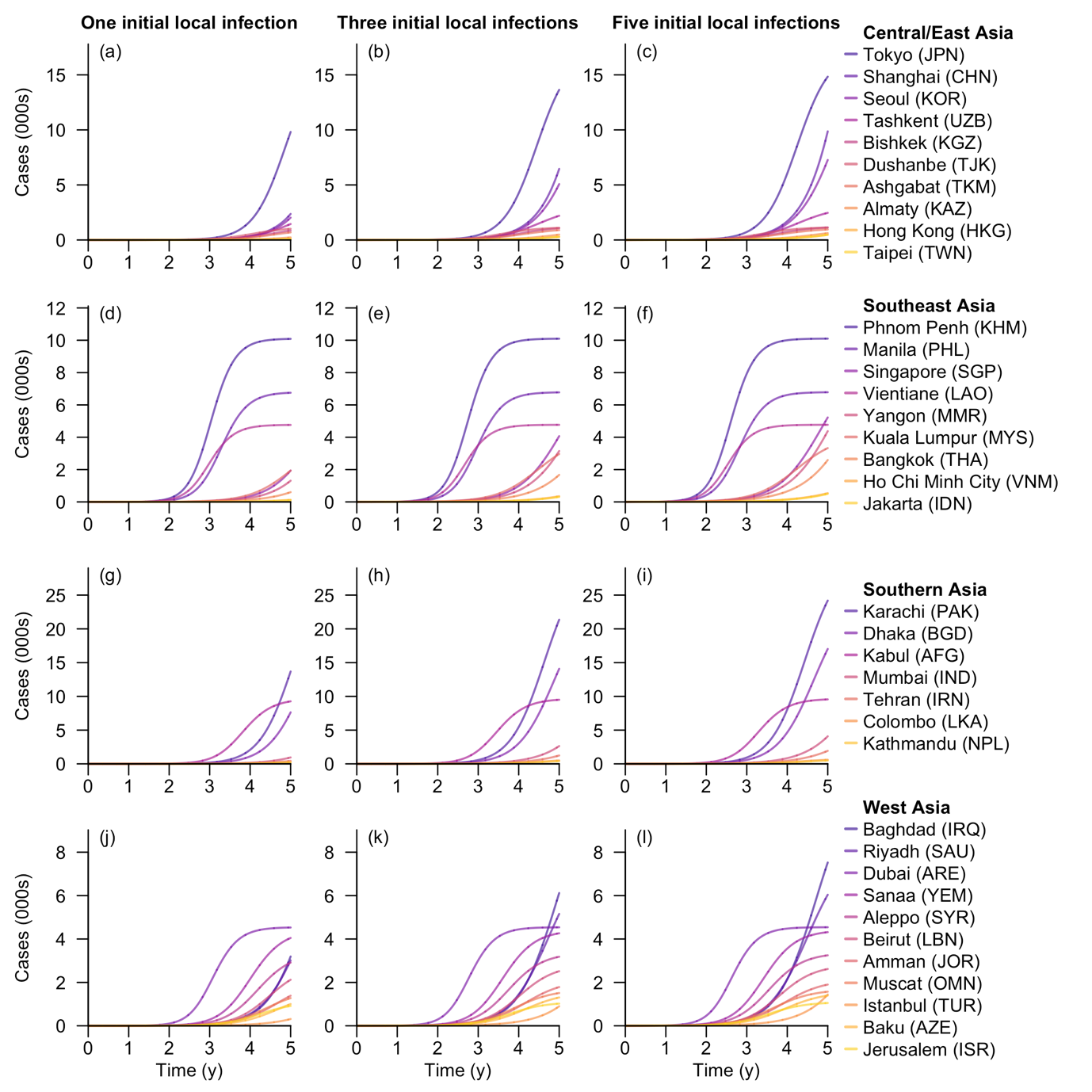
**Figure S2. Cumulative number of cases for each Asian city under the scenario with varying initial local infection sizes and a 14-day infectious period for undiagnosed infections.** Time (in years) refers to the duration since the initial local infections caused by a single importation event. The three columns represent scenarios with one, three, and five initial local infections, in which the epidemic curves were the average across settings with varying numbers of initials local infections in the low- and high-risk groups, respectively. Cities within each subregion were ordered by total number of cases in five years after five initial local infections.

### Model validation using the surveillance data in the DRC

We utilized our compartmental model to predict the death counts in South Kivu and compared with the surveillance data [16]. We set the overall population as seven million, with 2.7% belonging to the high-risk group. Proportion of susceptible individuals susceptible to the disease was 90.1% in the general population and 93.4% in the high-risk group [6]. We assumed a fixed importation rate of one per three days to account for spillover from other places and potential zoonotic transmission [11]. The simulated trajectory indicated the first case was likely to be detected in September 2023 (Figure S3, Table S3–S4), consistent with findings in literature [17].

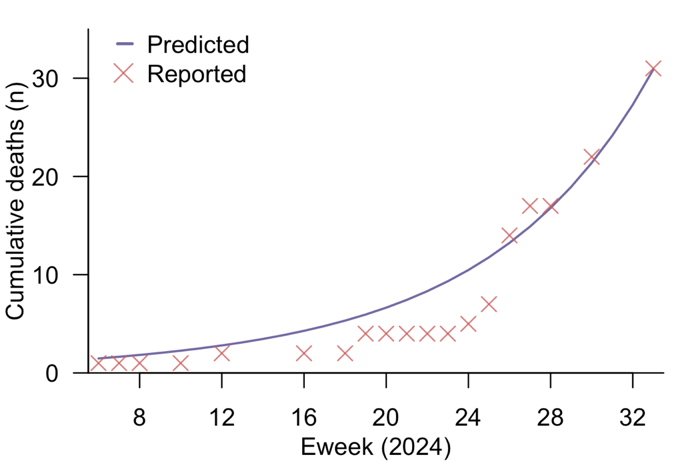
**Figure S3. Predicted cumulative number of deaths in South Kivu in 2024, assuming an importation rate of one per three days.** The blue line represents model predictions, while the red crosses are reported values in sitreps [11].

Considering the uncertainty surrounding the cause of the mpox outbreak in the DRC and the frequency of importation events, we further validated the model using the scenario aligning with our projections for Asian cities. Specifically, we assumed a single initial local infection within the high-risk population with no additional importation events and predicted the death counts in South Kivu, obtaining similar results (Figure S4, Table S3–S4).

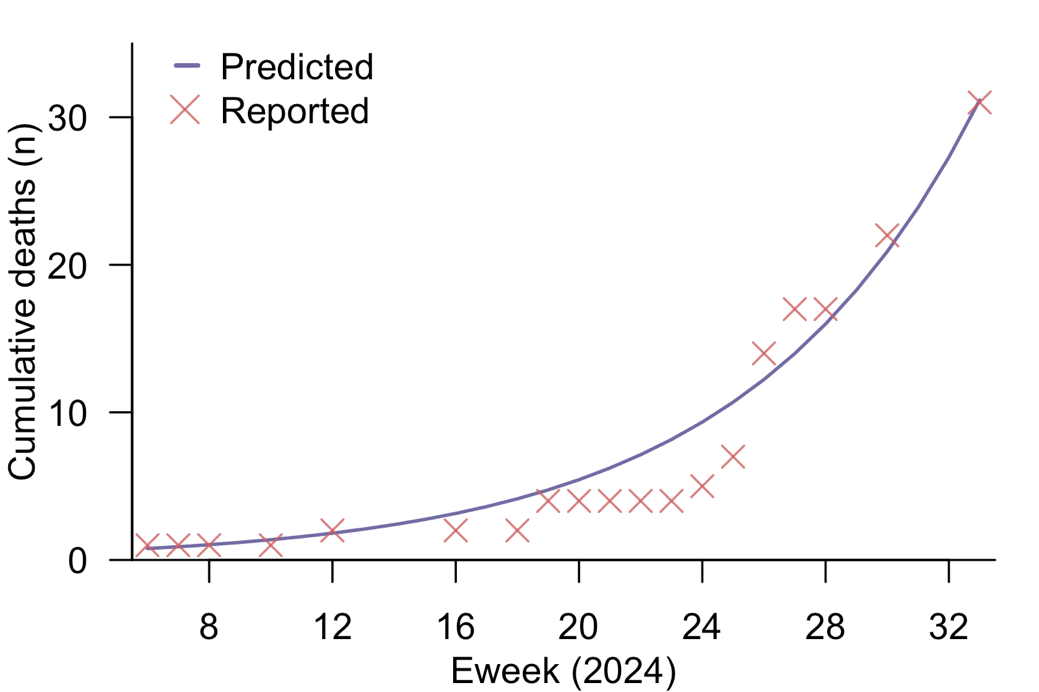

**Figure S4. Predicted cumulative number of deaths in South Kivu in 2024, assuming a single local infection in the high-risk group as the result of a one-time importation event.** The blue line represents model predictions, while the red crosses are reported values in sitreps [11].

**Table S3. Reported and predicted cumulative** **number of deaths in South Kivu in 2024, assuming either a constant importation rate of one per three days or a single importation event leading to one initial local infection in the high-risk group.** The death counts in column two are sourced from sitreps [11].

| **Eweek**  **(2024)** | **Reported**  **(from sitreps)** | **Predicted**  **(assuming continual importation)** | **Modelled**  **(assuming one initial local infection)** |
| --- | --- | --- | --- |
| 6 | 1 | 1.5 | 0.8 |
| 7 | 1 | 1.6 | 0.9 |
| 8 | 1 | 1.8 | 1.0 |
| 10 | 1 | 2.3 | 1.4 |
| 12 | 2 | 2.8 | 1.8 |
| 16 | 2 | 4.3 | 3.2 |
| 18 | 2 | 5.3 | 4.1 |
| 19 | 4 | 5.9 | 4.7 |
| 20 | 4 | 6.6 | 5.4 |
| 21 | 4 | 7.4 | 6.2 |
| 22 | 4 | 8.3 | 7.1 |
| 23 | 4 | 9.3 | 8.2 |
| 24 | 5 | 10.5 | 9.3 |
| 25 | 7 | 11.8 | 10.7 |
| 26 | 14 | 13.2 | 12.2 |
| 27 | 17 | 14.9 | 14.0 |
| 28 | 17 | 16.8 | 16.0 |
| 30 | 22 | 21.4 | 20.9 |
| 33 | 31 | 30.9 | 31.2 |

**Table S4. Reported and predicted cumulative** **number of cases in South Kivu in 2024, assuming either a constant importation rate of one per three days or a single importation event leading to one initial local infection in the high-risk group.** The death counts in column two are sourced from sitreps [11].

| **Eweek**  **(2024)** | **Reported**  **(from sitreps)** | **Predicted**  **(assuming continual importation)** | **Modelled**  **(assuming one initial local infection)** |
| --- | --- | --- | --- |
| 20 | 229 | 274 | 234 |
| 21 | 229 | 308 | 268 |
| 22 | 229 | 345 | 307 |
| 23 | 229 | 388 | 351 |
| 24 | 243 | 436 | 401 |
| 25 | 410 | 491 | 459 |
| 26 | 438 | 553 | 525 |
| 27 | 1010 | 623 | 600 |
| 28 | 977 | 703 | 686 |
| 30 | 1200 | 898 | 896 |
| 33 | 1370 | 1303 | 1335 |

### Sensitivity analysis: Impact of assuming an all-or-nothing smallpox vaccine to model effectiveness

Under the assumption that smallpox vaccines provide full protection against mpox infections for a fraction of the vaccinated population, we modelled the impacts of smallpox vaccination by excluding the corresponding population from the susceptible cohort (i.e., treating them as ‘recovered’). Let $R_{0}$ be the average number of secondary infections generated by an infectious individual in a fully susceptible population, $\gamma$ the recovery rate for diagnosed cases, and $N$ the total population size. The rate of infection was calculated as $\gamma R_{0}/N$.

Compared to the epidemic trajectories simulated in the main analysis, this modelling approach generated outbreaks with larger sizes in the first year following the initial local infections, but the overall affected populations were slightly smaller over the entire epidemic wave (Figure S5–S6).

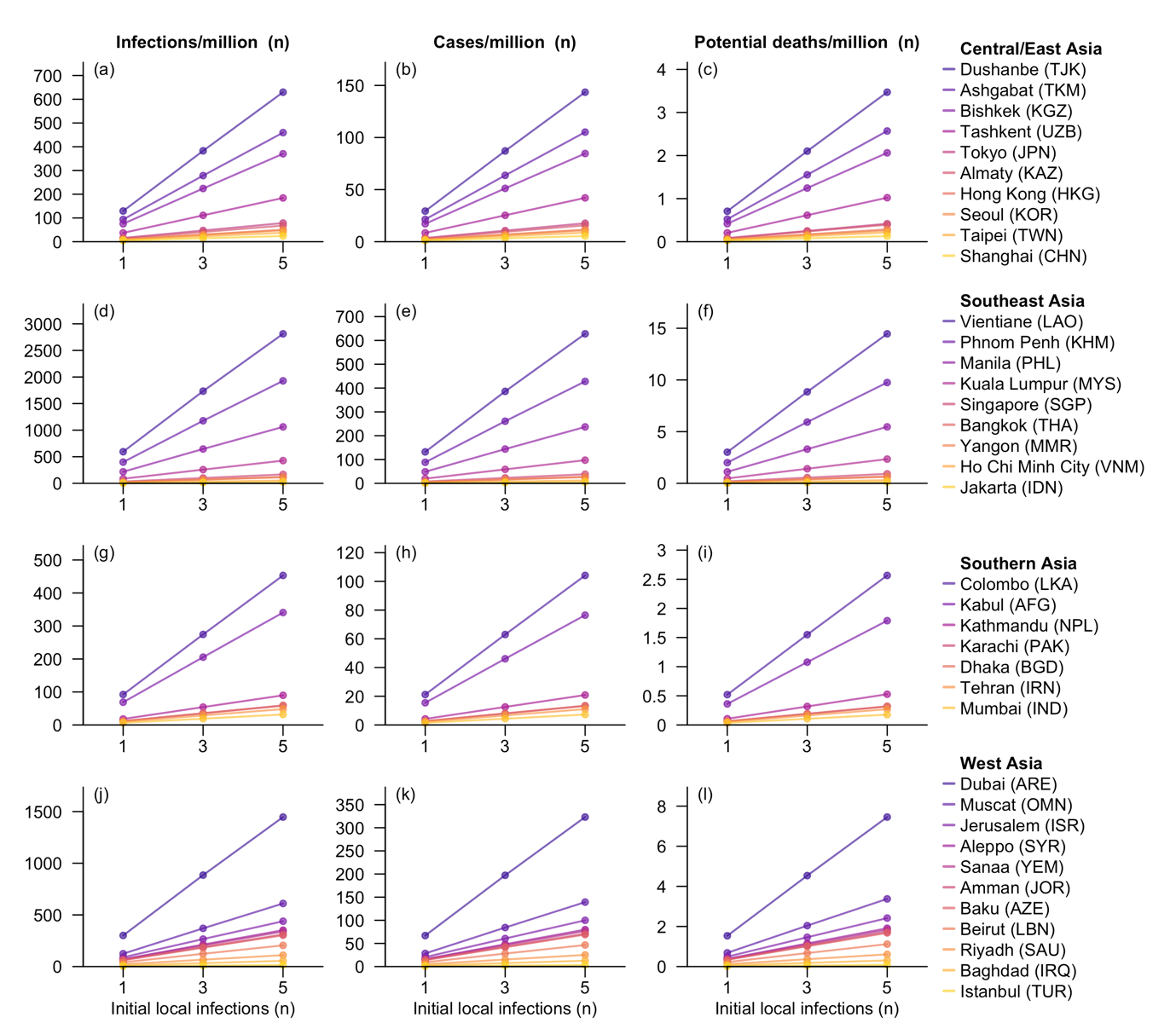
**Figure S5. Summary statistics for outbreaks in 37 Asian cities, assuming full protection provided by smallpox vaccines for a subset of the vaccinated populations.** The three statistics reported are: number of infections (Column 1), confirmed cases (Column 2), and individuals at risk of dying (Column 3) per million residents within one year following the initial local infections caused by a single importation event. The scenarios considered for each city include one, three, and five initial local infections. Cities within each subregion were ordered by potential death counts in the scenario with five initial local infections.

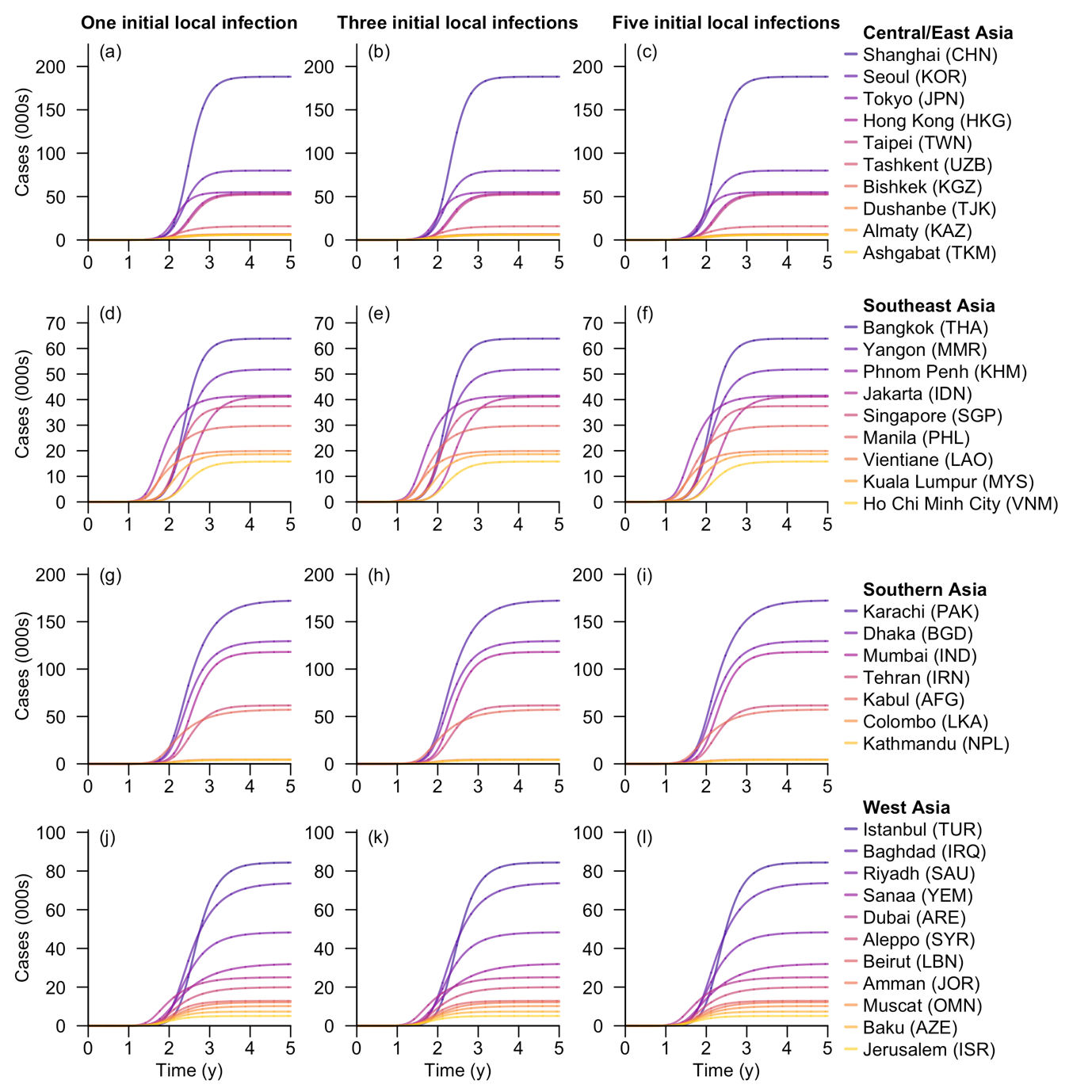
**Figure S6. Cumulative number of cases for each Asian city under the scenario with varying initial local infection sizes.** Vaccines were modelled to provide full protection for a subset of the vaccinated population. Time (in years) refers to the duration since the initial local infections caused by a single importation event. The three columns represent scenarios with one, three, and five initial local infections, in which the epidemic curves were the average across settings with varying numbers of initials local infections in the low- and high-risk groups, respectively. Cities within each subregion were ordered by total number of cases in five years after five initial local infections.

### Sensitivity analysis: Impact of varying high-risk subpopulation size on local outbreaks

In the main analysis, we estimated the size of the high-risk subpopulation from the number of sex workers and their clients using statistics from UNAIDS, IUSW, and Carael et al [18–20]. For territories lacking specific data, we approximated the figures using regional averages. To address the potential bias in the estimated population, we conducted a sensitivity analysis, assuming that the high-risk group accounted for 2%, 5%, or 10% of individuals aged 15–49 years in each city or territory. We simulated epidemic curves for each city under scenarios involving one, three, or five initial local infections. Great disparities were observed in the proportion of susceptible individuals in the 15–49 age group across the 37 territories, with this group accounting for 40–60% of individuals in most territories (Figure S7). The simulation results are visualized in Figure S8–S10. The ranking of outbreak sizes for the 37 cities was similar across the three high-risk subpopulation proportions, but substantially differed from the main analysis, in which we assumed heterogenous proportions of high-risk individuals among people aged 15–49 years in each city (Figure 4).

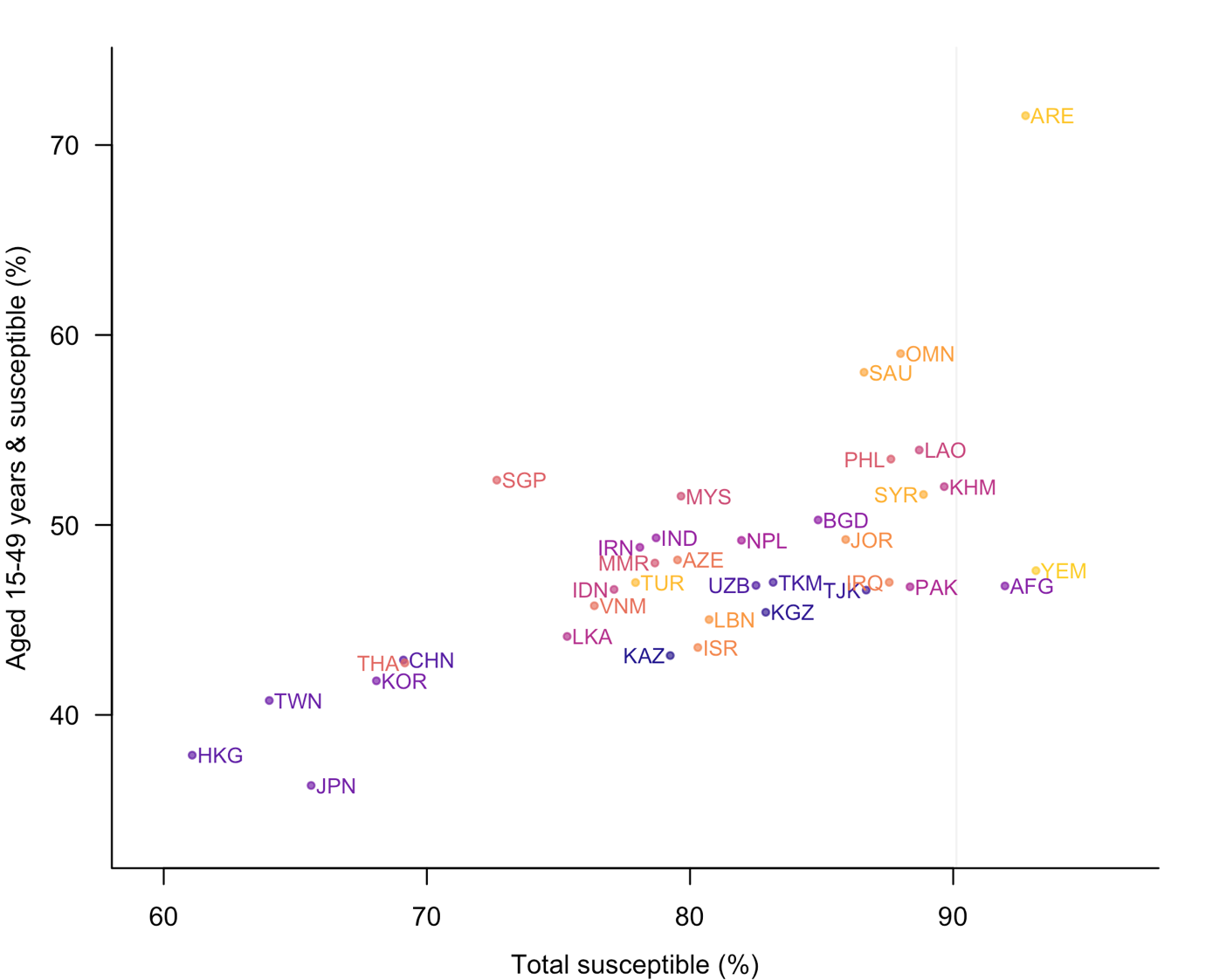
**Figure S7. Proportion of total susceptible population (x-axis) and that of susceptible individuals aged 14–49 years (y-axis) in each territory.** A vaccine efficacy of 80.7% was assumed for smallpox vaccines. Territories within the same subregion (Central, East, Southeast, Southern, or West Asia) are represented by similar colours. The grey vertical line indicates the overall proportion of susceptible population in the DRC.

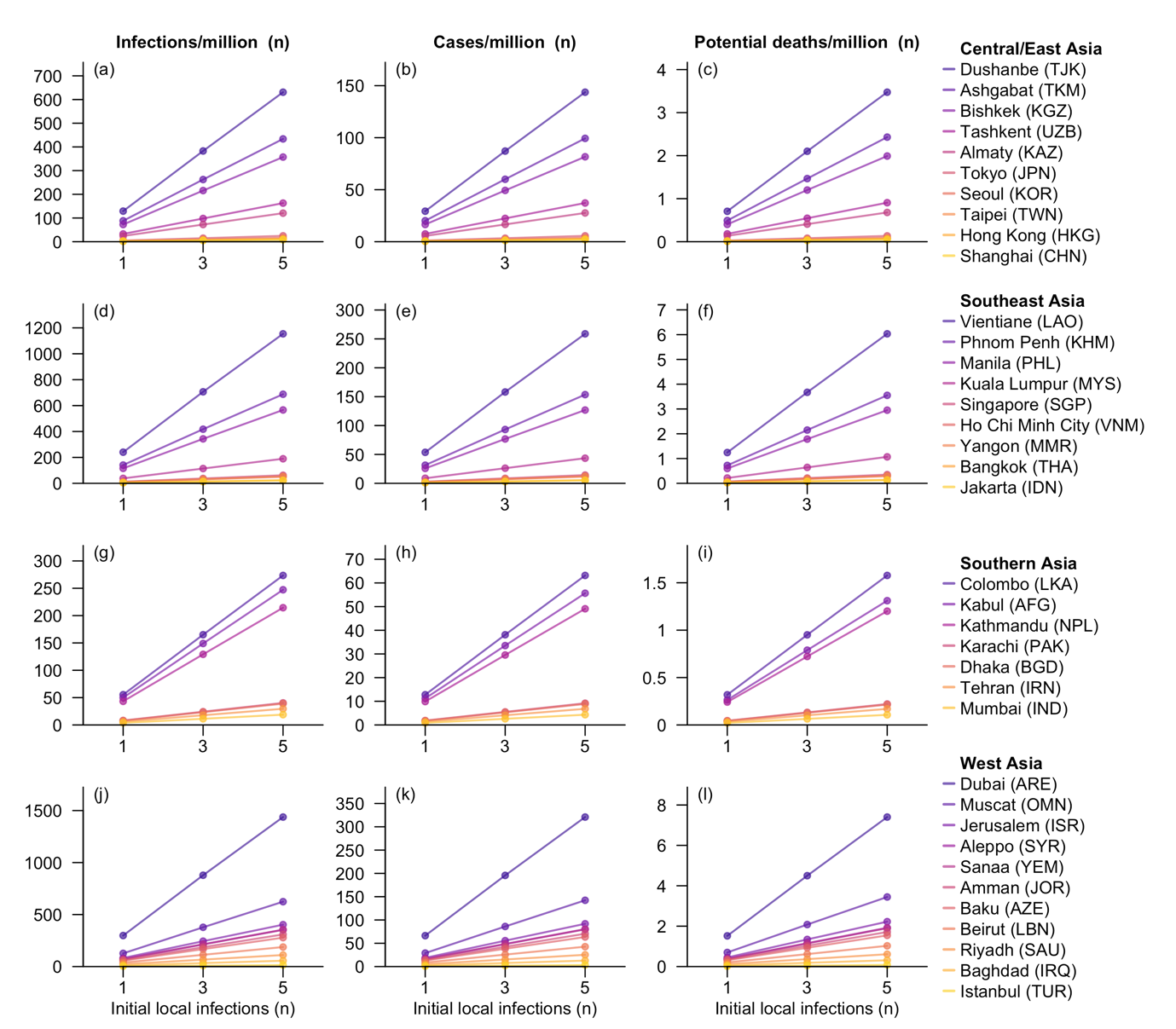
**Figure S8. Summary statistics for outbreaks in 37 Asian cities, assuming that 2% of individuals aged 15–49 years comprised the high-risk subpopulation.** The three statistics reported are: number of infections (Column 1), confirmed cases (Column 2), and individuals at risk of dying (Column 3) per million residents within one year following the initial local infections caused by a single importation event. The scenarios considered for each city include one, three, and five initial local infections. Cities within each subregion were ordered by potential death counts in the scenario with five initial local infections.

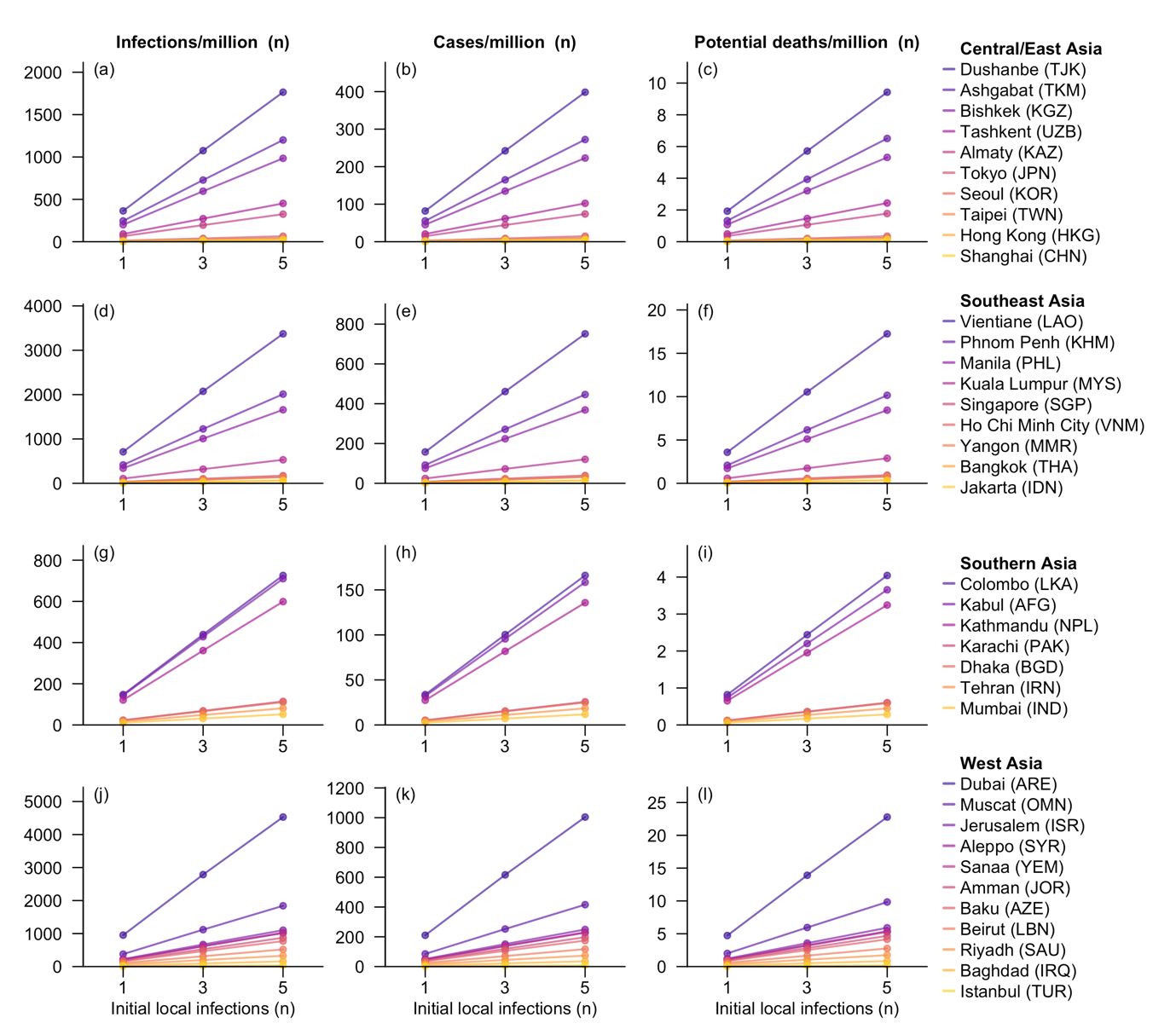
**Figure S9. Summary statistics for outbreaks in 37 Asian cities, assuming that 5% of individuals aged 15–49 years comprised the high-risk subpopulation.** The three statistics reported are: number of infections (Column 1), confirmed cases (Column 2), and individuals at risk of dying (Column 3) per million residents within one year following the initial local infections caused by a single importation event. The scenarios considered for each city include one, three, and five initial local infections. Cities within each subregion were ordered by potential death counts in the scenario with five initial local infections.

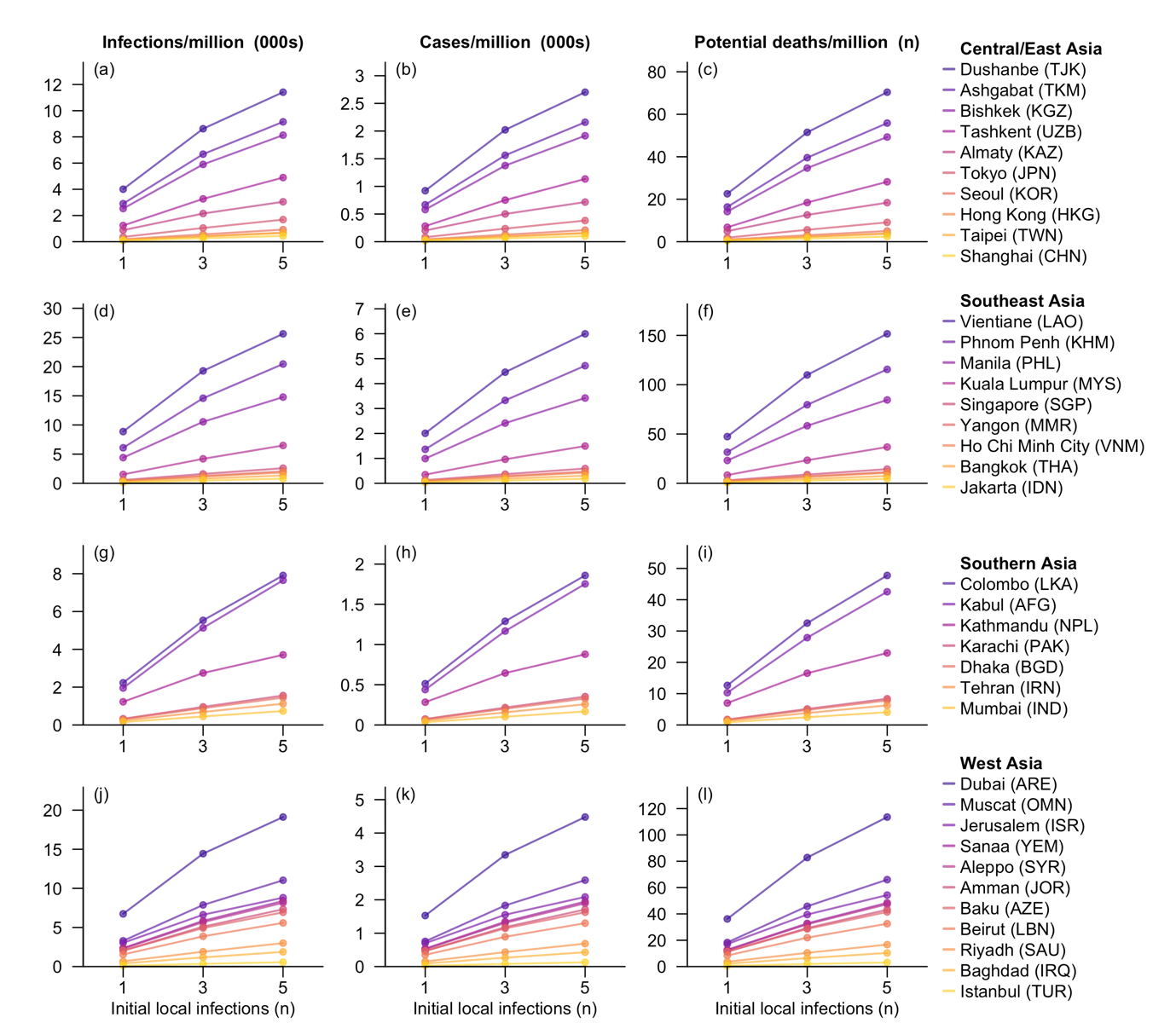
**Figure S10. Summary statistics for outbreaks in 37 Asian cities, assuming that 10% of individuals aged 15–49 years comprised the high-risk subpopulation.** The three statistics reported are: number of infections (Column 1), confirmed cases (Column 2), and individuals at risk of dying (Column 3) per million residents within one year following the initial local infections caused by a single importation event. The scenarios considered for each city include one, three, and five initial local infections. Cities within each subregion were ordered by potential death counts in the scenario with five initial local infections.

### Sensitivity analysis: Impact of initial local infections contracting the disease through sexual contact

In the main analysis, we assumed that initial local infections contracted the disease through community contact, resulting in the probability of these infections occurring within the high-risk group being equal to the prevalence of high-risk individuals in the overall population. Nevertheless, this probability would significantly increase if at least one initial local infection was infected through sexual contact. Therefore, we explored an extreme scenario in which all initial local infections were high-risk individuals.

Under this scenario, the epidemic trajectories exhibited significantly larger outbreak sizes over one year compared to those in the main analysis, regardless of the initial number of local infections. This indicates faster outbreak dynamics. Additionally, the non-linear relationship between the initial local infection size and the total number of infections over one year in some cities suggests a potential deceleration in outbreak growth rate by the end of the first year. However, the total affected populations over the entire epidemic wave remained the same as those in the main analysis (Figure S11–S12).

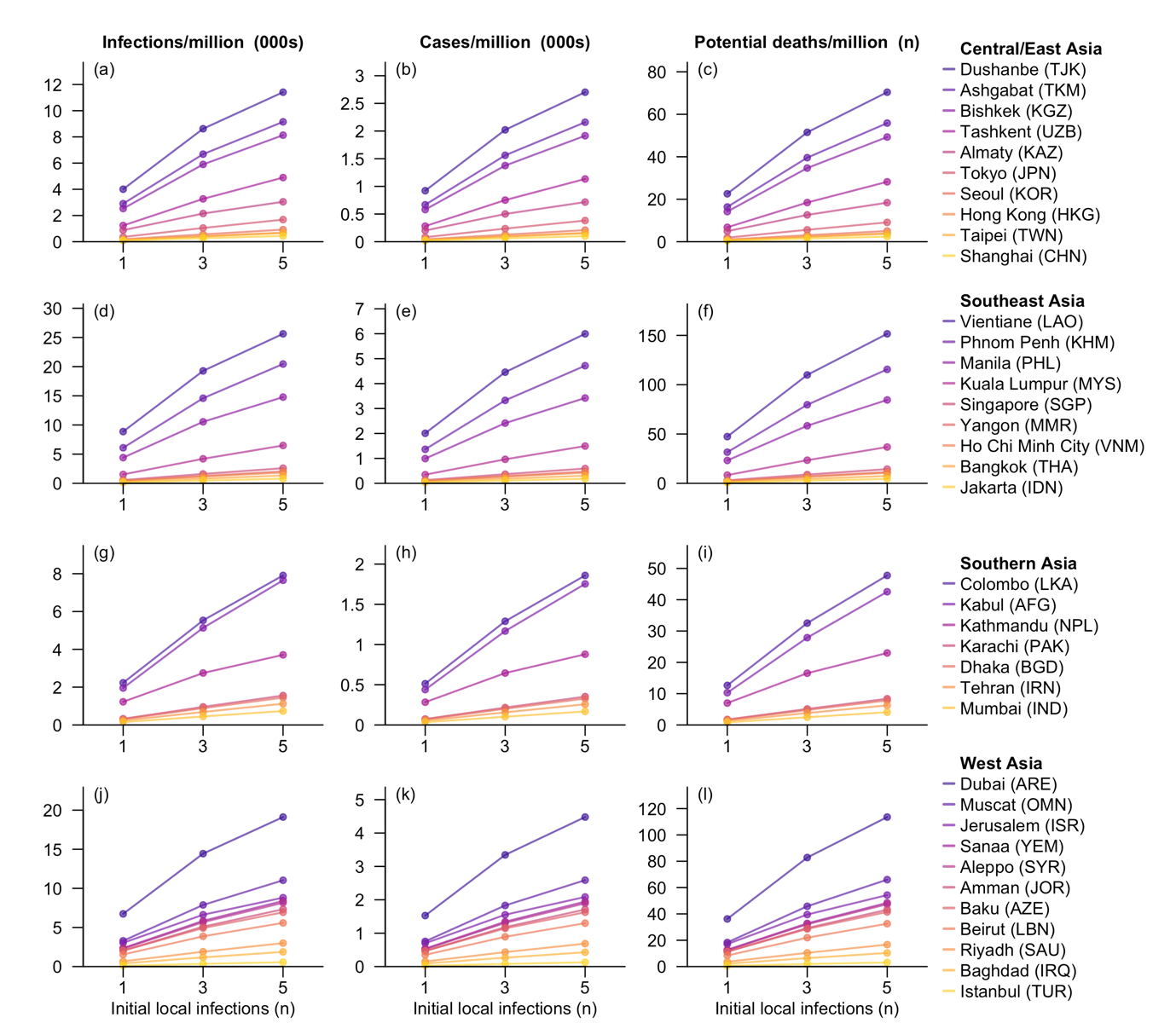
**Figure S11. Summary statistics for outbreaks in 37 Asian cities, assuming all initial infections belonging to the high-risk group.** The three statistics reported are: number of infections (Column 1), confirmed cases (Column 2), and individuals at risk of dying (Column 3) per million residents within one year following the initial local infections caused by a single importation event. The scenarios considered for each city include one, three, and five initial local infections. Cities within each subregion were ordered by potential death counts in the scenario with five initial local infections.

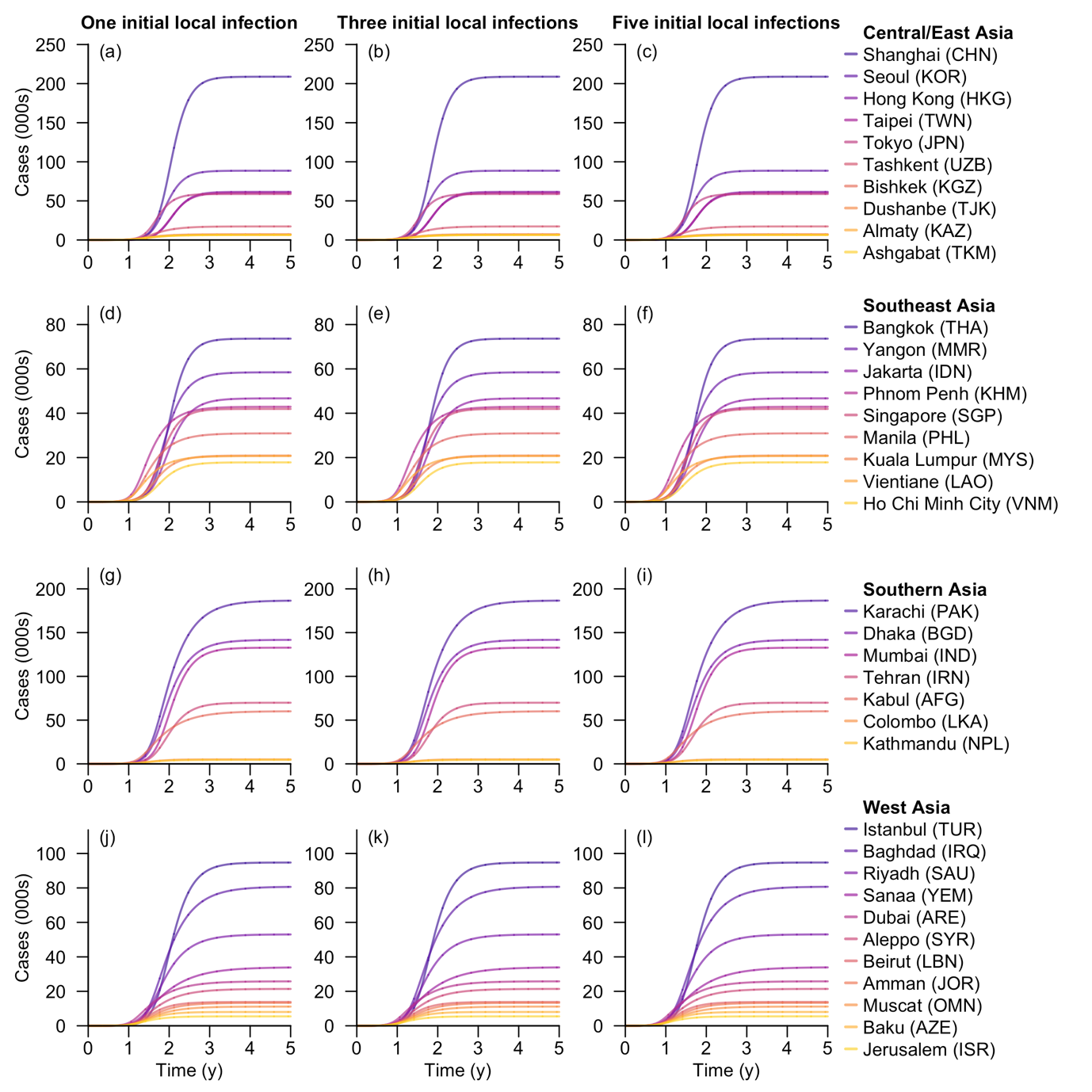
**Figure S12. Cumulative number of cases for each Asian city under the scenario with varying initial local infection sizes.** All the initial infections were assumed to belong to the high-risk group. Time (in years) refers to the duration since the initial local infections caused by a single importation event. The three columns represent scenarios with one, three, and five initial local infections, in which the epidemic curves were the average across settings with varying numbers of initials local infections in the low- and high-risk groups, respectively. Cities within each subregion were ordered by total number of cases in five years after five initial local infections.

We further simulated outbreaks in each city assuming fewer initial local infections being high-risk compared to the aforementioned extreme scenario, and visualized in Figure S13 the number of infections per thousand residents in the first year for each scenario and city. A non-linear increase would be expected with the increase in the proportion of high-risk individuals among the initial local infections.

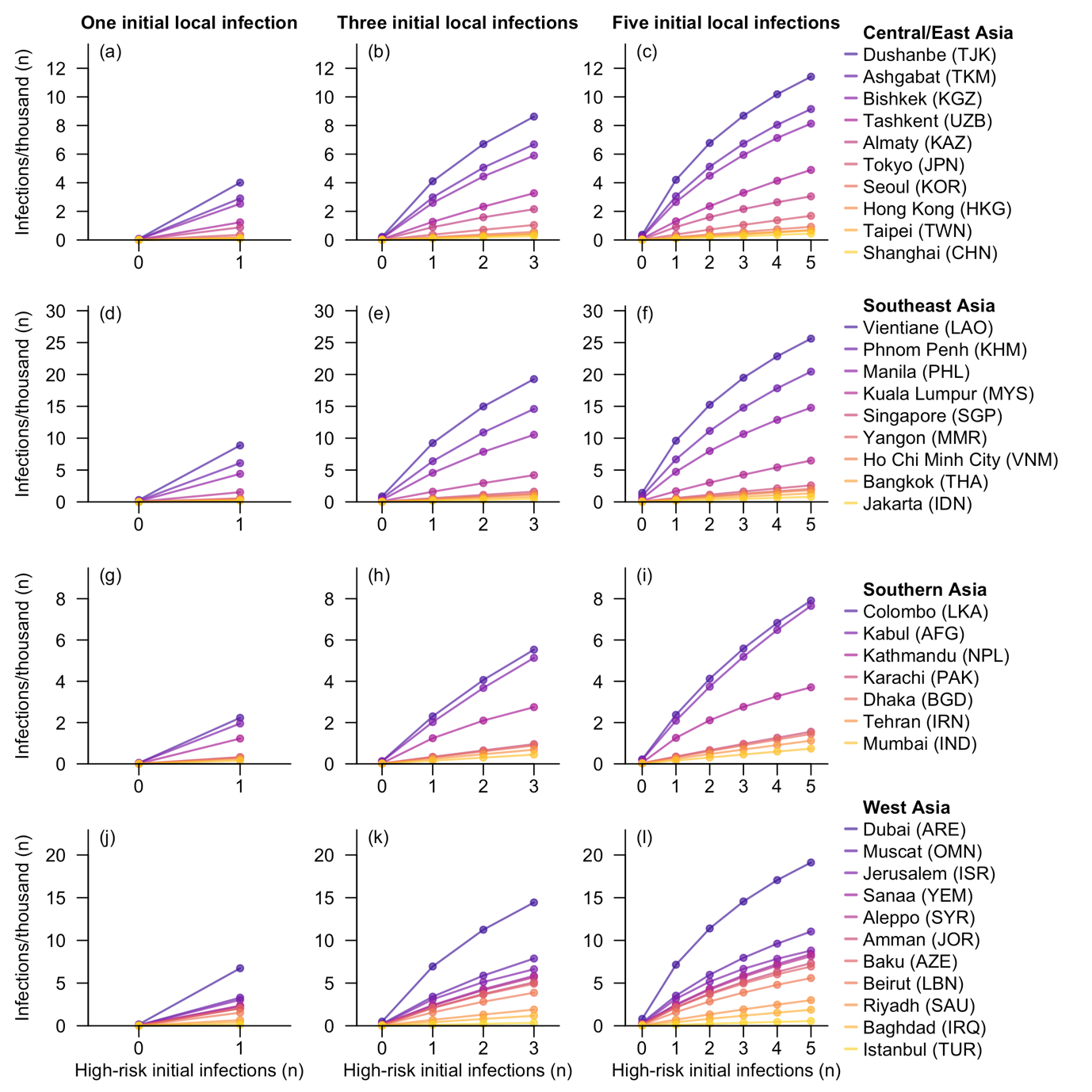
**Figure S13. Outbreak size in 37 Asian cities, assuming varying numbers of initial infections belonging to the high-risk group. This is measured by number of infections per thousand residents** within one year following the initial local infections caused by a single importation event. The scenarios considered for each city include one, three, and five initial local infections, corresponding to the three columns in the figure. Cities within each subregion were ordered by outbreak size in the scenario with five initial local high-risk infections.

### Additional simulation results

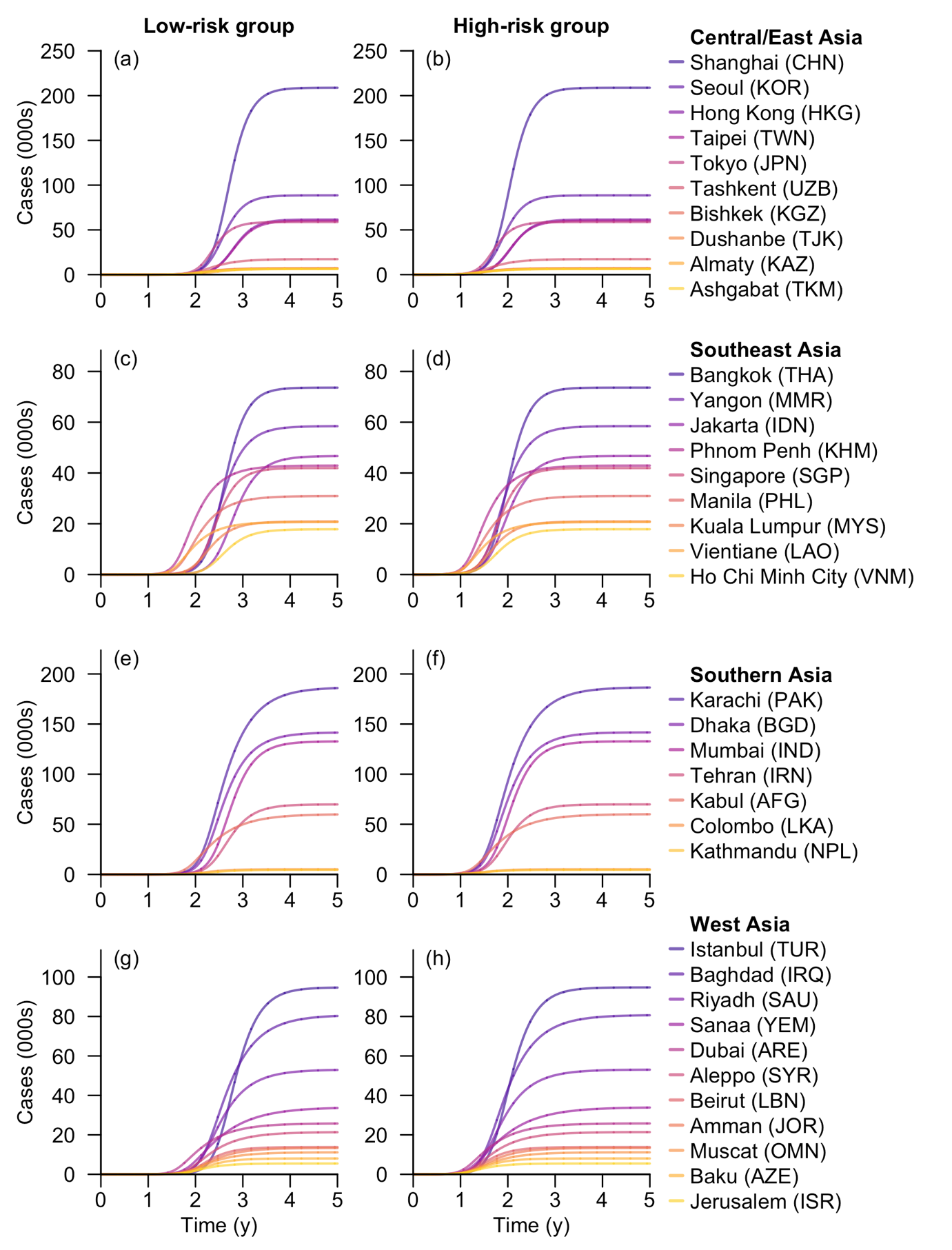
**Figure S14. Cumulative number of cases over five years for each Asian city under the scenario with one initial local infection.** Time (in years) refers to the duration since the initial local infections caused by a single importation event. The two columns represent the index local infection occurring in the low- or high-risk group, respectively. Cities within each subregion were ordered by total number of cases in four years after the initial local infection in the high-risk group.

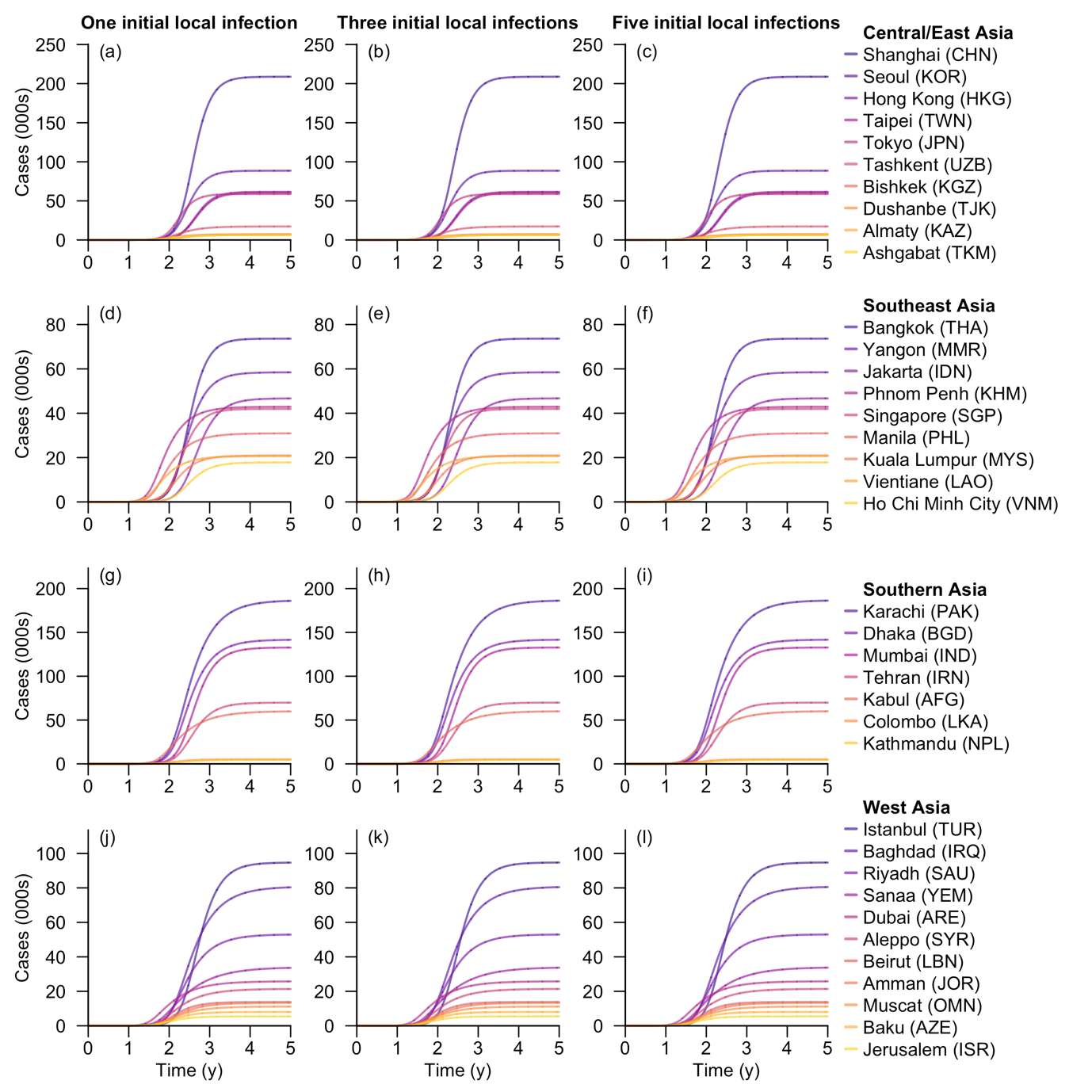
**Figure S15. Cumulative number of cases for each Asian city under the scenario with varying initial local infection sizes.** Time (in years) refers to the duration since the initial local infections caused by a single importation event. The three columns represent scenarios with one, three, and five initial local infections, in which the epidemic curves were the average across settings with varying numbers of initials local infections in the low- and high-risk groups, respectively. Cities within each subregion were ordered by total number of cases in five years after five initial infections.

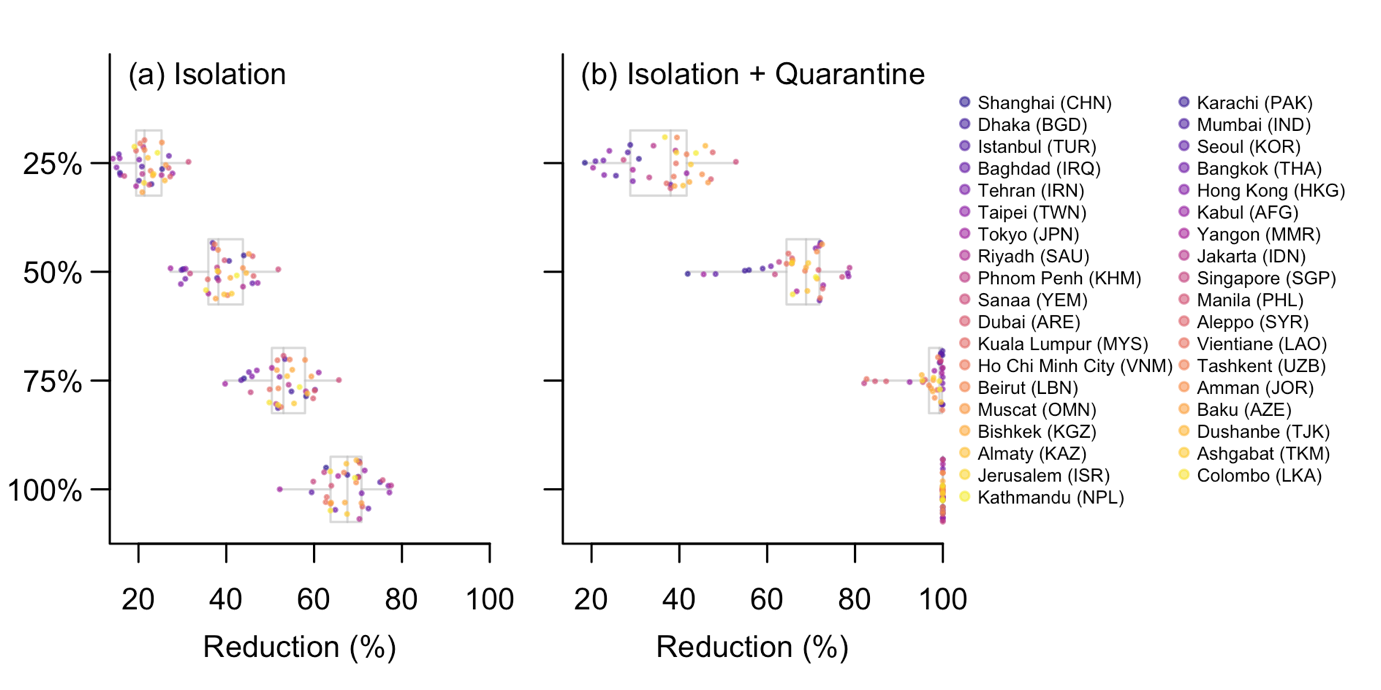
**Figure S16. Projected reduction in confirmed cases in five years following the initial three local infections due to (a) isolation or (b) isolation combined with quarantine.** The reduction proportions were calculated for each level of isolation effectiveness (25%, 50%, 75%, and 100% reduction in interpersonal contacts). compared to the baseline scenario with no NPIs and three initial local infections per city. Grey box plots display the median, interquartile ranges, and ranges of reductions across the 37 cities. In the legend bar on the right, these cities were ordered by confirmed case counts in five years in the scenario without NPIs, with a darker colour indicating a greater decrease.

**Table S5. Summary of intervention effectiveness.** The summary statistics include medians and interquartile ranges [IQRs] for projected reductions in confirmed cases in five years following the initial three local infections due to NPIs across the 37 Asian cities. The unit ‘pp’ stands for percentage point.

| **Isolation effectiveness (%)** | **Reduction due to isolation (pp)** | **Reduction due to quarantine (pp)** |
| --- | --- | --- |
| 25 | 21.4 (19.5–25.3) | 15.5 (8.3–18.9) |
| 50 | 38.2 (35.9–43.8) | 27.6 (25.7–30.0) |
| 75 | 53.0 (50.4–57.9) | 44.7 (39.4–48.9) |
| 100 | 67.6 (63.7–70.8) | 32.4 (29.2–36.2) |

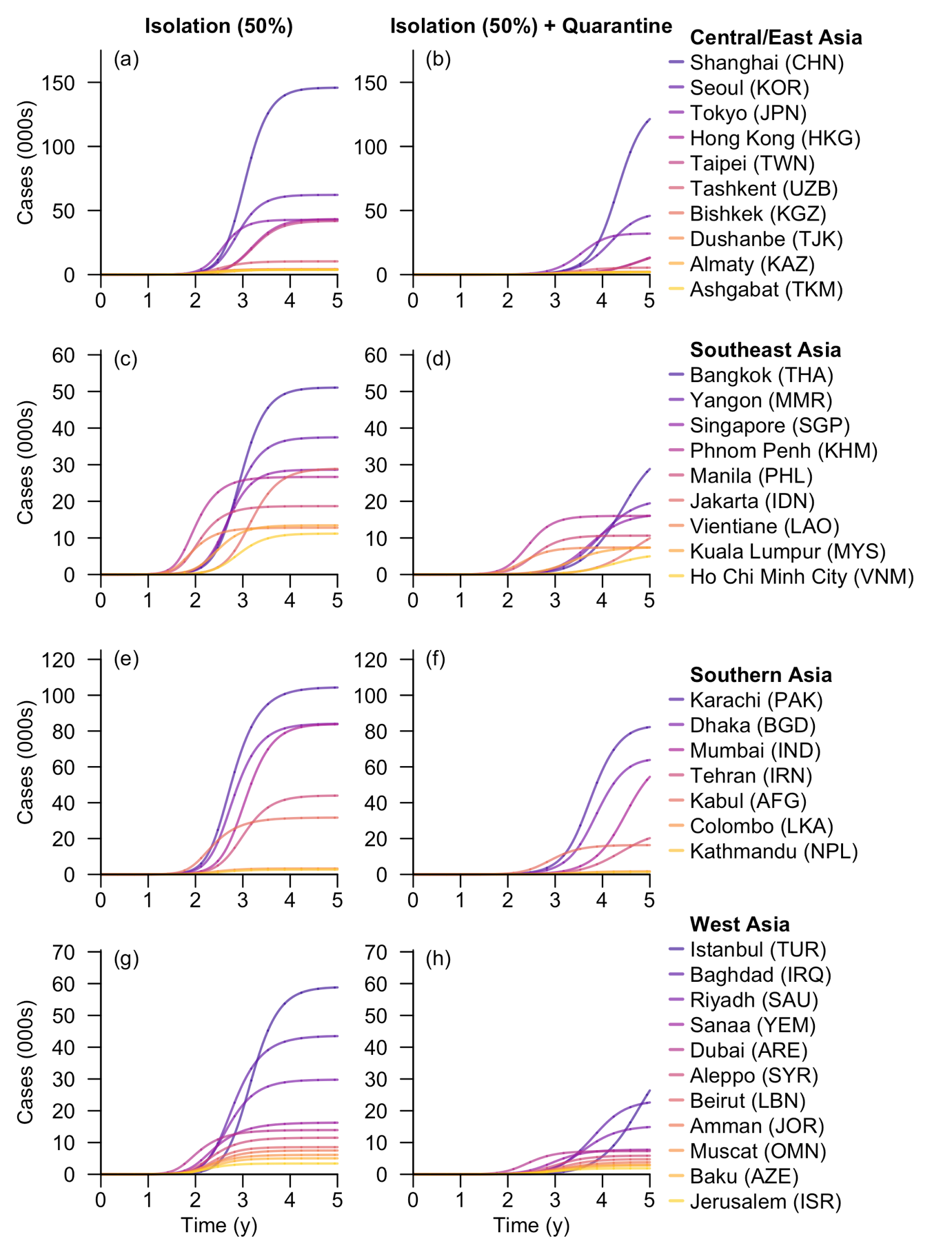
**Figure S17.** **Cumulative number of cases for each Asian city under the scenarios with interventions.** Time (in years) refers to the duration since the initial local infections caused by a single importation event. The two columns represent scenarios without and with the implementation of quarantine. In both scenarios, the outbreak started with three initial local infections and diagnosed cases were isolated with 50% effectiveness. The epidemic curves were the average across settings with varying numbers of initials local infections in the low- and high-risk groups. Cities within each subregion were ordered by total number of cases in five years after five initial local infections.

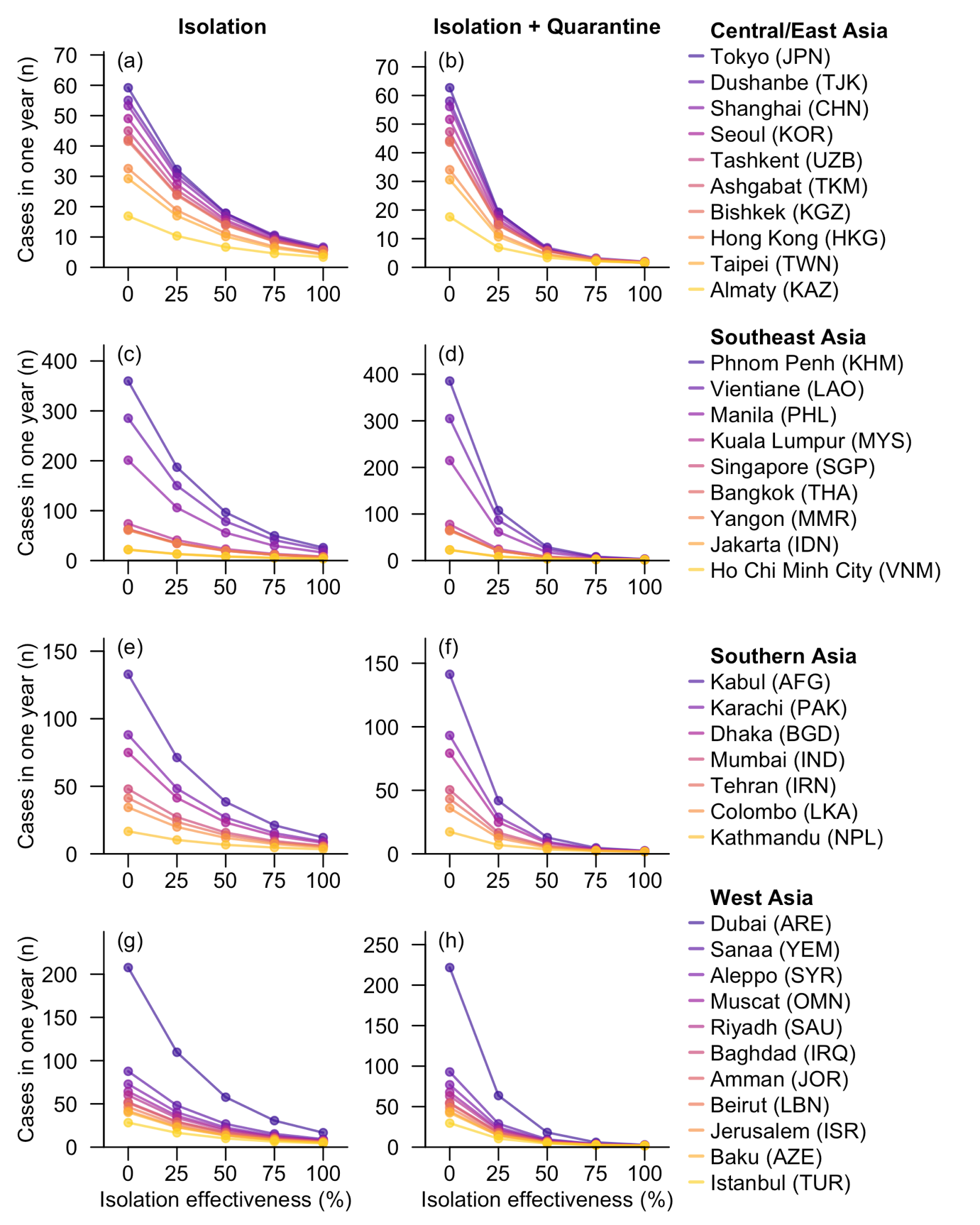
**Figure S18. Confirmed cases in one year following the initial three local infections under different intervention scenarios.** These include the baseline scenario with no intervention and scenarios with varying levels of isolation effectiveness (25%, 50%, 75%, and 100% reduction in interpersonal contacts). Columns 1 and 2 represent the NPI strategies of isolation only and isolation combined with quarantine, respectively. Cities within each subregion were ordered by confirmed case counts in one year in the scenario without NPIs.

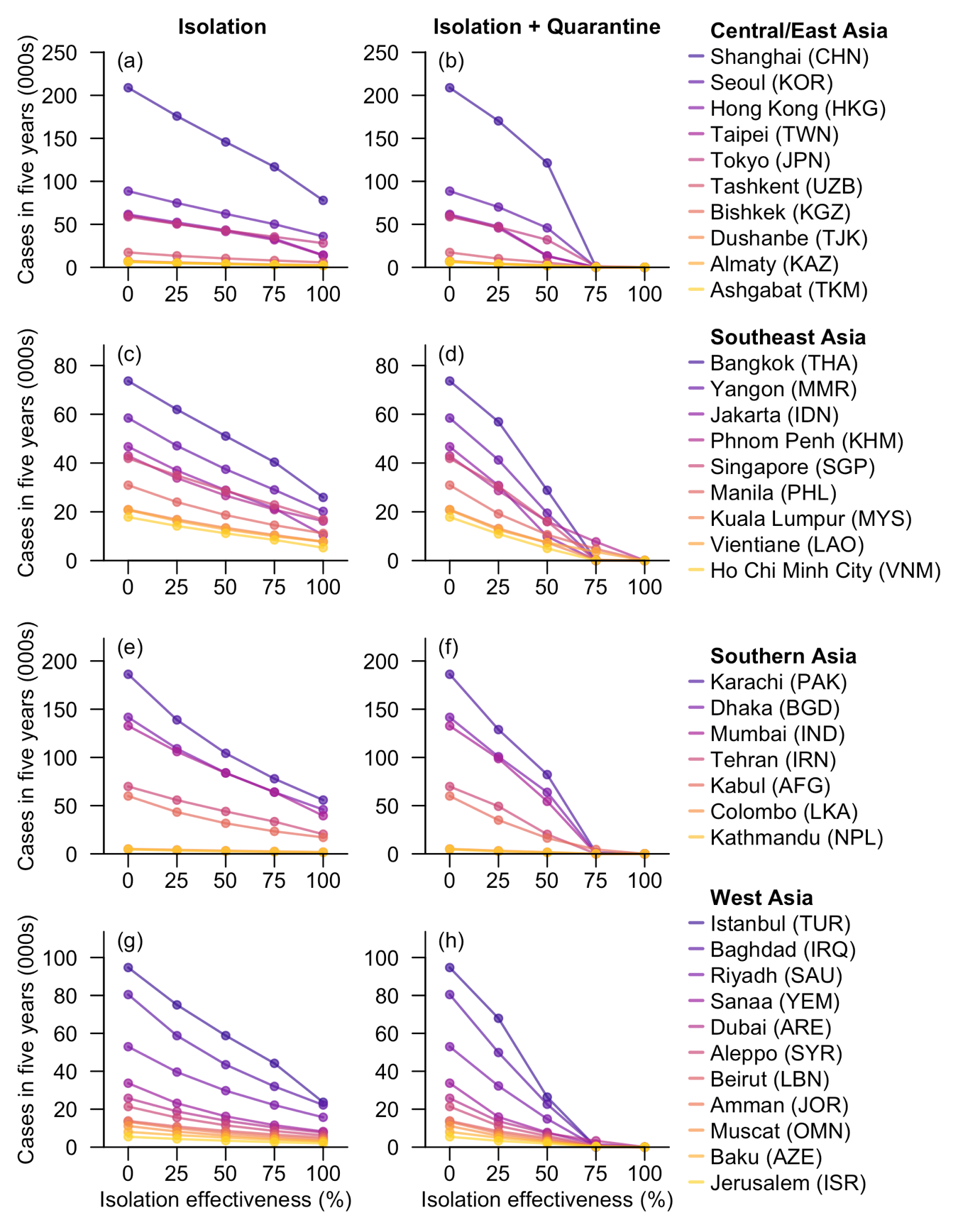
**Figure S19. Confirmed cases in five years following the initial three local infections under different intervention scenarios.** These include the baseline scenario with no intervention and scenarios with varying levels of isolation effectiveness (25%, 50%, 75%, and 100% reduction in interpersonal contacts). Columns 1 and 2 represent the NPI strategies of isolation only and isolation combined with quarantine, respectively. Cities within each subregion were ordered by confirmed case counts in five years in the scenario without NPIs.
